# Improving the accuracy of Respiratory Syncytial Virus (RSV) incidence among hospitalised adults in Bristol, UK

**DOI:** 10.1101/2025.01.24.25321075

**Authors:** Katie Lihou, Robert Challen, Anastasia Chatzilena, George Qian, Glenda Oben, Jade King, Serena McGuinness, Begonia Morales-Aza, Kaltun Duale, Ainhoa Rodriguez Pereira, William Healy, Jennifer Oliver, Nick Maskell, Adam Finn, Leon Danon, Catherine Hyams, the Avon CAP Research Group

**Author notes:** **Corresponding Author:** Catherine Hyams, MB PhD, Bristol Vaccine Centre, University of Bristol, St Michael’s Hill, Bristol BS2 8AE, United Kingdom.

## Abstract

Respiratory Syncytial Virus (RSV) burden in adults is of interest due to recently-licensed vaccines, however, estimates are affected by test characteristics.

We conducted a prospective cohort study of adults with acute lower respiratory tract disease (aLRTD) hospitalised in Bristol from April 2022–March 2023. RSV was detected by RT-PCR at standard-of-care and by additional nasopharyngeal, saliva, and sputum samples. Latent class analysis quantified and adjusted for test error and multiple testing.

6906/11445 aLRTD cases (60%) were tested and 251 were positive (3.6%). Test-positivity peaked in December (7.9-12.7%). Among cases, 43% had pneumonia and 55% had non-pneumonic infection. Due to low positivity-rates and imperfect specificity, test-positivity (3.6%) overestimated true prevalence (2.3%). Adjusted adult population incidence/1000-person-years was 0.33 (0.21-0.49), and 2.02 (1.10-3.06) in ≥75-year-olds.

RSV vaccines could reduce morbidity of hospitalised adult aLRTD, including non-pneumonic infection. Adult RSV burden accuracy is improved by adjustment for test characteristics, particularly estimates out-of-season.

## INTRODUCTION

Respiratory syncytial virus (RSV) is a recognised cause of paediatric illness [1] and its importance in adult disease, including exacerbation of underlying heart and lung disease, is increasingly recognised [2–3]. RSV severity in adults varies widely, from mild symptoms to hospitalisation and even death, particularly in older or frailer individuals [4]. A recent meta-analysis in high-income countries, pre-pandemic, found the pooled hospitalisation rate for RSV-related illness in ≥65-year-olds/1000 population/year was 1.57, and 3.47, after simple adjustment for diagnostic test sensitivity [5]. New RSV vaccines have been approved to prevent respiratory disease in older adults [6–8]. Therefore, it is crucial to have accurate adult RSV-related disease burden estimates to assess the potential impact and cost-effectiveness of RSV vaccines and inform future public health policy.

Few studies report RSV burden following SARS-CoV-2 emergence [9]. Social distancing measures implemented to reduce COVID-19 [10–12] affected the circulation of other respiratory pathogens. No prospective UK studies have reported the disease burden of RSV ARI hospitalisations among adults. Three retrospective pre-pandemic time-series modelling studies estimated disease burden among British older adults by associating the variability in an RSV-indicator (i.e., RSV laboratory surveillance data) with outcomes that included RSV-related events (i.e., respiratory hospitalisations). These UK studies reported an average annual incidence of 1.56/1000 population in >65y [13] and 2.65/1000 in those with high-risk medical conditions; an annual incidence of 2.51/1000 respiratory hospital admissions [≥75y; 2010–2017] and 0.71/1000 respiratory hospital admissions [65–74y] [14]; and, average annual incidences of 0.90/1000 respiratory hospital admissions [65-74y], 2.80/1000 respiratory hospital admissions [75-84y] and 6.0/1000 respiratory hospital admissions [≥85y] [15].

Published RSV-related incidence among adults has varied substantially, driven by differences in methodology, underlying RSV prevalence, and testing-rates and characteristics. RSV testing in adults is infrequent due lack of RSV-specific treatments [16]. Clinical suspicion for RSV-associated cardiopulmonary disease exacerbation tends to be low and, consequently, these patients are often not tested [3]. Additionally, routine testing-rates of ARI patients decreases with patient age [17]. Other under-ascertainment sources include using case definitions that exclude some RSV disease presentations (e.g. pneumonia only), reduced sensitivity of single specimen PCR testing among older adults [18–19], sampling period misaligning with RSV season, and delays between illness onset and sampling [20].

Test error rates act as a source of potential over- or under-estimation of disease burden estimates. However, test error is more likely to result in overestimation of RSV-related hospitalisation burden in adults as population incidence is generally low [21]. In low prevalence populations, imperfect test specificity, the true negative rate, can have a larger impact on disease burden estimates than imperfect test sensitivity [22]. As the relative proportion of disease-negative individuals is high, false positive-rates (1-specificity) result in a high ratio of false positives to true positives. Therefore, the proportion of test-positives correctly identified (positive predictive values (PPV)), will be low relative to the test negatives correctly identified (negative predictive values (NPV);[23–24]). This is particularly problematic in study designs implementing multiple testing in parallel and classifying any positive test result as a positive individual, since combined test specificity decreases with increasing numbers of tests [25].

Realised test error rates are impacted by test sensitivity and specificity, as well as variations in the testing process, such as test settings, sample quality and volume, and patient characteristics. Latent class analysis (LCA) quantifies relative test accuracy within a specific system, even in the absence of a gold-standard reference test, incorporating factors impacting the false positive and false negative rate, including test specificity and sensitivity [26–28]. LCA classifies the probability of individuals being disease positive or negative, which are unobserved latent classes, based on the results of multiple imperfect tests that are combined to form a composite reference standard. LCA can quantify the effect of multiple testing on disease diagnosis certainty, and this method has been used to quantify test accuracy in the absence of a gold standard reference test for multiple pathogens [28–33]. Therefore, LCA can be used to adjust for study-specific test characteristics to improve the accuracy of disease burden estimation.

This analysis reports RSV test-positivity rate, by subgroup and over time, among adults in two large hospitals serving the population of Bristol, UK, using a prospective, population-based active surveillance study. LCA is used to adjust for test error rates and multiple testing in estimations of true RSV prevalence and population incidence, an error source commonly ignored in disease burden estimations [34]. The primary objective is to determine the RSV disease burden in this population following SARS-CoV-2 emergence and report incidence and its associated uncertainty.

## METHODS

### Study design

This prospective observational cohort study included adults admitted to two large university hospitals (Southmead, and Bristol Royal Infirmary) in Bristol, UK. Adults (≥18y) admitted to both hospitals from 1st April 2022–31st March 2023, encompassing all acute secondary care in Bristol, were screened for study inclusion. This time-period was selected as it included the first winter season unaffected by COVID-19 pandemic-related measures. Inclusion and exclusion criteria were designed to capture all aLRTD (acute lower respiratory tract disease) cases as previously published [35].

Demographic and clinical data were collected from medical records systematically using REDCap [36]. We collected data on co-morbidities at admission, determining Charlson co-morbidity index (CCI;[37]) and Rockwood clinical frailty score (score 5–9 indicating frailty;[38]). Vaccination records were obtained from linked general practitioner (GP) records. All cause 30-day mortality numbers were obtained by linkage with the national healthcare records using a unique patient identifier.

### Respiratory specimen testing

Standard-of-care (SOC) virological testing results obtained from naso- or oropharyngeal (NP/OP) swabs were taken from patients’ medical charts. SOC samples underwent RSV testing by RT-PCR in the local UKHSA clinical microbiology lab using one of three different panel tests run on either Hologic Panther Fusion platform or BioFire Diagnostics system. Patients who consented to participate in the enhanced diagnostic testing study arm underwent research sampling between April 2022–March 2023, providing one or more of the following specimen types: an upper respiratory swab (NP or combined NP/OP swab), a saliva, or sputum sample. If subjects were unable to produce saliva, a saline mouth wash specimen was collected. Research specimens were tested by RT-PCR using Certest Biotech Viasure® viral pathogen multiplex panels. The sample volumes, PCR threshold and baseline values used, differed between SOC and research RT-PCR tests. Research swabs were stored in STGG (skim-milk, tryptone, glucose, glycerine) medium and SOC swabs in VTM (viral transport medium).

### Case definitions

aLRTD was defined as any presentation with acute lower respiratory disease, including pneumonia; non-pneumonic-LRTI (NP-LRTI); acute bronchitis; exacerbations of underlying cardiorespiratory disease (CRDE) including chronic obstructive pulmonary disease (COPD) and asthma; and acute or decompensated heart failure (HF). Patients were designated RSV test-positive if they tested positive on PCR, from any specimen type. For full case definitions see: Extended Data, 1;[39–40]).

### Statistical analysis

The primary objectives were to report RSV test-positivity rates among the RSV tested aLRTD hospitalised population and sub-categories, and estimate population incidence rates with appropriate uncertainty, accounting for test error and multiple testing (Fig. 1). Comparisons between different participant populations were made using Kolmogorov–Smirnov tests for continuous variables, and Fisher’s exact test for categorical variables. Test-positivity rates were calculated as the number of RSV-positives/100 aLRTD hospital admissions tested for RSV. Results are stratified by age and aLRTD subgroups. Apart from pneumonia and NP-LRTI, these subgroups are not mutually exclusive. RSV test-positive-rates over time were estimated using the weekly number of test-positives and the weekly number of individuals tested for RSV in the aLTRD admitted population, fitted to a quasi-binomial model assuming a time-varying rate. Data were locally fitted with order 2 polynomial and a logit link function using the methods of [41]. All analyses were conducted using R [42].

**Figure 1:**
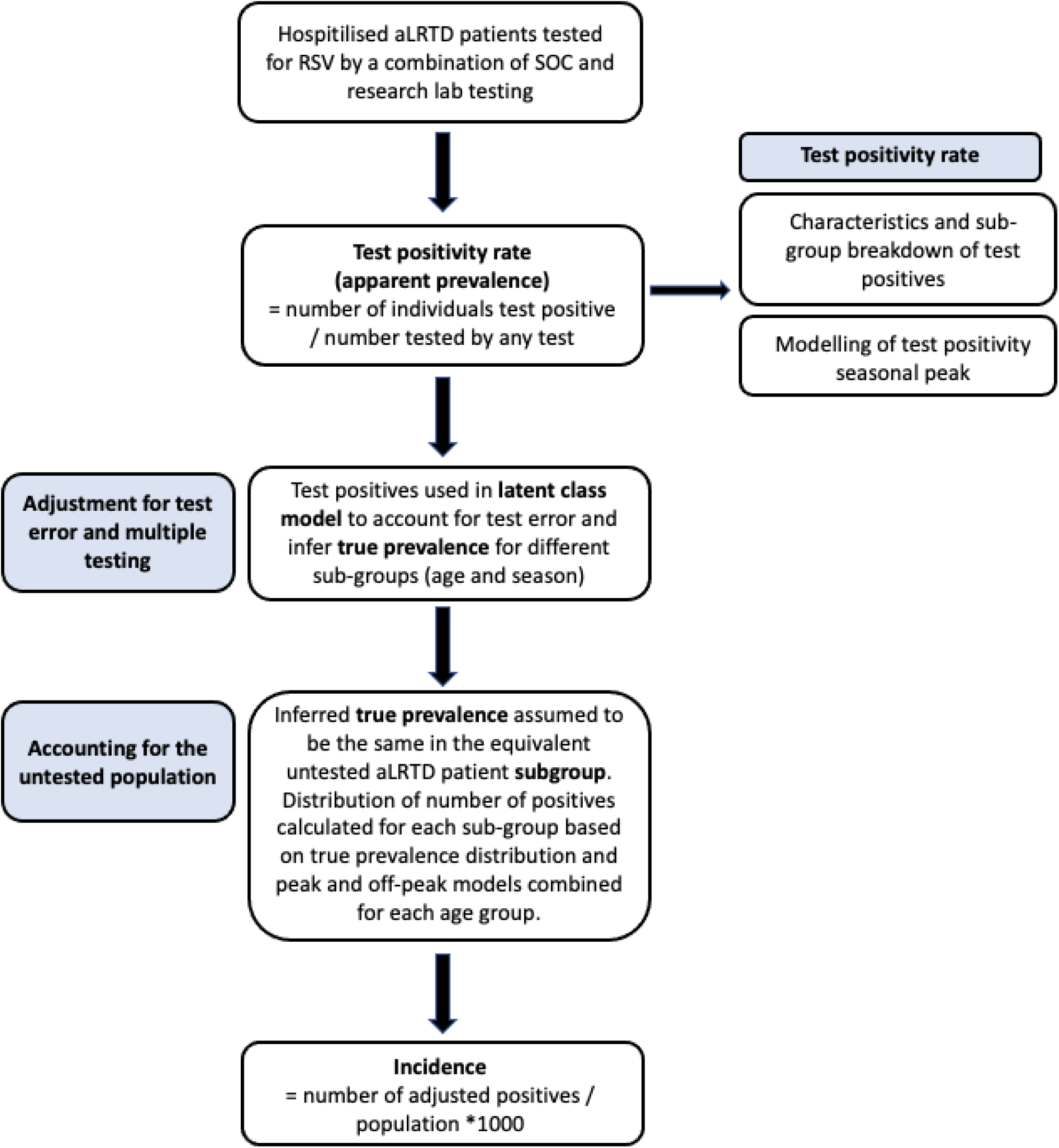
Flowchart of methods used to calculate annual RSV population incidence in adults in Bristol between April 2022 – March 2023.

### Latent class analysis (LCA)

Latent class analysis (LCA) was used to adjust prevalence estimates in the tested population for test error and multiple testing. The LC model was built using a Bayesian framework and implemented in Stan using the No-U-Turn sampler variant of Hamiltonian Monte Carlo through the R interface Rstan [43]. The probability of an individual having a particular pattern of test outcomes is conditional on disease prevalence and test sensitivity and specificity, which are unobserved latent parameters estimated by the model (for details see Supplementary). Sensitivity is the probability that the test will be positive if an individual is infected with RSV (Se = *P*(Test + | Infected)), and specificity is the probability that the test will be negative if an individual is not infected with RSV (Sp = *P*(Test - | Not infected)); the model requires no assumptions about these as the agreement and disagreement between multiple tests provides enough information to infer them. LC models assume all tests are conditionally independent of one another, therefore an individual varying random effect with a scaling parameter was included to model the conditional dependence between the three research sample types [30;44-45].

Model accuracy was confirmed using simulated data with known parameters of prevalence, test sensitivity, and test specificity. Test error associated with multiple testing was quantified by combining individual inferred test sensitivities and specificities (see Supplementary).

### Population incidence

The LC model was run for different subgroups of the RSV-tested aLRTD population: the peak RSV season (Nov-Feb), the off-peak season and the peak season split by age group. Incidence was calculated in each age group by assuming the same inferred true prevalence in the tested and untested subgroup populations (Number of RSV-positives = inferred prevalence * number of aLRTD hospitalised patients in subgroup). The number of positives in each age group was based on the median from the combined distributions of the number of positives from peak/off-peak models. Uncertainty in the number of positives was based on 95% credible intervals calculated using the package ‘bayestestR’ (method=HDI;[46]). These numbers were used to calculate the adult population RSV incidence/1000-person-years with appropriate uncertainty (Fig. 1).

## RESULTS

### Characteristics of RSV testing

Between 1^st^ April 2022 and 31^st^ March 2023, 118,819 adults were admitted at the two study sites: 11,445 (10%) had aLRTD. 6,906 (60%) were tested for RSV (Extended Data, 2;3): 3,632 (53%) were tested by standard-of-care (SOC); 1,633 (24%) by research sampling; and 1,641 (24%) using both approaches (Extended Data, 4). Median time from admission to first SOC test was 0d (days; IQR:0-2), and to research test was 2d (IQR:1-3; Extended Data, 5). Testing-rate varied throughout the year, peaking over the winter due to increased SOC testing (Fig. 2). Overall, 39.7% (4539/11,445) of participants and during peak season (Nov-Feb), 26.2% (900/3,432) were untested. The median age of the tested participants was 73.4y (IQR:60.3-82.5), and this differed somewhat between participants by testing-type (Extended Data, 4), with the biggest difference in ≥85-year-olds: 23.1% of SOC-only-tested were ≥85y compared to 16.8% of research-only-tested. RSV-tested participants were slightly younger than untested participants (Extended Data, 3). Among those tested, 7.2% resided in care homes, the median CCI score was 4.0 (IQR:3.0-6.0), and 61.3% were smokers/ex-smokers. Tested participants were more likely to have pneumonia and CRDE than untested participants (47.0% versus 37.4%, and 51.2% versus 40.9%, respectively; Extended Data, 3).

**Figure 2:**
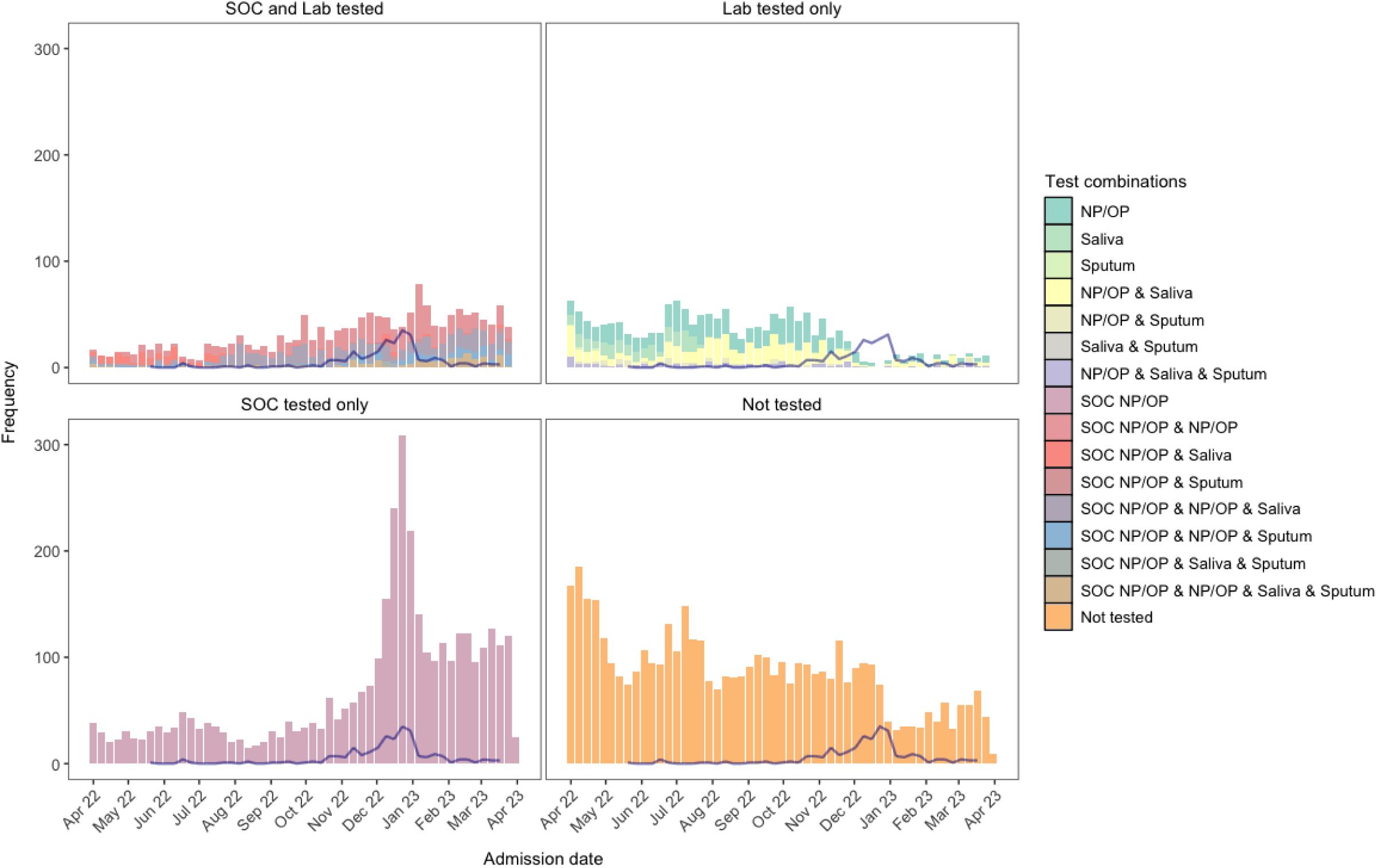
Test and sample type combinations for testing of RSV across the study time period. Histogram of specimen combinations collected from each study participant (between April 2022 – March 2023); SOC NP/OP swab; research NP/OP swab; research saliva sample; and research sputum sample. Weekly counts of RSV, by a positive on any test, are shown by the blue line.

Of participants undergoing RSV testing, 4.4% (95%CI:3.9-5.0%) tested positive on SOC testing alone and 3.6% (3.2-4.1) through any test (i.e., SOC NP/OP; research NP/OP, saliva, or sputum): 232/251 (92.4%) RSV test-positive participants were identified through SOC testing (Table 1). The percentage of RSV test-positive individuals varied across individuals with different combinations of tests (Fig. 5). Of the research samples, the RSV test-positive-rate was 1.1% (0.7-1.5) for NP/OP, 0.9% (0.5-1.5) for saliva, and 3.4% (2.0-5.3) for sputum samples (Table 1).

**Table 1:** Test-positive-rates of the different RSV tests in the study. The number tested, percentage tested, number positive, and percentage positive of different types of test/samples for RSV in the study. 95% CIs are Clopper and Pearson.

| Study Specimens | n, tested | %, tested with specimen type | n, positive by specimen type | %, positive of sample tested | 95% CI |
| --- | --- | --- | --- | --- | --- |
| Overall | 6906 | 100.0% | 251 | 3.6% | 3.2 – 4.1% |
| SOC NP/OP swab | 5273 | 76.4% | 232 | 4.4% | 3.9 – 5.0% |
| Research lab, any sample | 3250 | 47.1% | 47 | 1.4% | 1.1 – 1.9% |
| NP/OP swab | 2887 | 41.8% | 31 | 1.1% | 0.7 – 1.5% |
| Saliva* | 1629 | 23.6% | 15 | 0.9% | 0.5 – 1.5% |
| Sputum | 530 | 7.7% | 18 | 3.4% | 2.0 – 5.3% |

\*In subjects that were unable to produce saliva, a saline mouth wash was obtained (1.9%; 31/1629).

### Characteristics of RSV-positive participants

Of the 251 RSV test-positive participants, 109 (43%) had pneumonia, 137 (55%) NP-LRTI, 149 (59%) CRDE, and 40 (16%) HF. 4.2% (149/3535) of CRDE patients and 3.2% (40/1265) of HF patients, that were tested for RSV, tested positive for RSV. RSV test-positives were found in all age groups, including 18-34-year-olds, while 75-84-year-olds had the highest percentage of RSV test-positives (Table 2). All RSV test-positive patients aged 18-34y had comorbid disease (data not shown). Among RSV test-positive participants, 5 (2%) also had a positive COVID-19 test, 6 (2%) a positive influenza-A test, and 1 (0%) a positive influenza-B test. The median length of hospital admission was 5d (IQR:2-8). Overall, 3.6% of RSV test-positive patients were admitted to ICU, remaining there for an average of 6d (IQR:3-14). The all-cause 30-day mortality rate was 4.4% and was highest among ≥75-year-olds (7.1%), broadly increasing with patient age (Table 3).

**Table 2:** Patient characteristics of adults hospitalised with aLRTD that were test-positive for RSV by standard of care (SOC) or research lab. Patient demographics are shown for all adults hospitalised with aLRTD during the study period that had an RSV-positive test from any test (total, pneumonia, NP-LRTI, CRDE, HF). Full demographics by RSV testing are presented in Extended Data, 3;4.

|  |  | Confirmed RSV infection |  |  |  |  |  |  |  |  |  |
| --- | --- | --- | --- | --- | --- | --- | --- | --- | --- | --- | --- |
| Variable | Characteristic | Total |  | Pneumonia |  | NP-LRTI |  | CRDE |  | HF |  |
|  |  | N | Value | N | Value | N | Value | N | Value | N | Value |
| Age | Median (IQR) | 251 | 75.0<br>(61.6-82.4) | 109 | 77.5<br>(64.7-82.7) | 137 | 73.2<br>(60.0-82.2) | 149 | 73.1<br>(63.0-80.7) | 40 | 79.9<br>(64.7-85.7) |
| CCI | Median (IQR) | 251 | 4.0 (3.0-6.0) | 109 | 4.0 (3.0-6.0) | 137 | 4.0 (3.0-5.0) | 149 | 4.0 (3.0-6.0) | 40 | 5.22 (40) |
| Age category (y) † | 18-34 | 10 | 4.0% | 4 | 3.7% | 5 | 3.6% | 6 | 4.0% | 1 | 2.5% |
|  | 35-49 | 23 | 9.2% | 5 | 4.6% | 16 | 11.7% | 12 | 8.1% | 1 | 2.5% |
|  | 50-64 | 43 | 17.1% | 19 | 17.4% | 24 | 17.5% | 25 | 16.8% | 9 | 22.5% |
|  | 65-74 | 48 | 19.1% | 21 | 19.3% | 27 | 19.7% | 38 | 25.5% | 3 | 7.5% |
|  | 75-84 | 81 | 32.3% | 39 | 35.8% | 40 | 29.2% | 50 | 33.6% | 15 | 37.5% |
|  | ≥85 | 46 | 18.3% | 21 | 19.3% | 25 | 18.2% | 18 | 12.1% | 11 | 27.5% |
| Gender | Male | 122 | 48.6% | 52 | 47.7% | 68 | 49.6% | 67 | 45.0% | 20 | 50.0% |
|  | Female | 129 | 51.4% | 57 | 52.3% | 69 | 50.4% | 82 | 55.0% | 20 | 50.0% |
| Pneumonia | yes | 109 | 43.4% | 109 | 100.0% | 0 | 0.0% | 65 | 43.6% | 21 | 52.5% |
| NP-LRTI | yes | 137 | 54.6% | 0 | 0.0% | 137 | 100.0% | 79 | 53.0% | 19 | 47.5% |
| CRDE | yes | 149 | 59.4% | 65 | 59.6% | 79 | 57.7% | 149 | 100.0% | 23 | 57.5% |
| HF | yes | 40 | 15.9% | 21 | 19.3% | 19 | 13.9% | 23 | 15.4% | 40 | 100.0% |
| Ethnicity | White British | 183 | 72.9% | 76 | 69.7% | 102 | 74.5% | 109 | 73.2% | 28 | 70.0% |
|  | White other | 7 | 2.8% | 4 | 3.7% | 3 | 2.2% | 5 | 3.4% | 2 | 5.0% |
|  | Black | 3 | 1.2% | 1 | 0.9% | 2 | 1.5% | 0 | 0.0% | 0 | 0.0% |
|  | Asian | 5 | 2.0% | 1 | 0.9% | 4 | 2.9% | 3 | 2.0% | 0 | 0.0% |
|  | Mixed origin | 3 | 1.2% | 0 | 0.0% | 3 | 2.2% | 0 | 0.0% | 2 | 5.0% |
|  | Other | 2 | 0.8% | 2 | 1.8% | 0 | 0.0% | 2 | 1.3% | 0 | 0.0% |
|  | Unknown | 48 | 19.1% | 25 | 22.9% | 23 | 16.8% | 30 | 20.1% | 8 | 20.0% |
| Care home resident | no | 232 | 92.4% | 98 | 89.9% | 129 | 94.2% | 141 | 94.6% | 37 | 92.5% |
|  | Yes | 19 | 7.6% | 11 | 10.1% | 8 | 5.8% | 8 | 5.4% | 3 | 7.5% |
| Smoker | Current | 35 | 13.9% | 17 | 15.6% | 18 | 13.1% | 24 | 16.1% | 5 | 12.5% |
|  | Ex-smoker | 117 | 46.6% | 52 | 47.7% | 62 | 45.3% | 77 | 51.7% | 18 | 45.0% |
|  | Non-smoker | 78 | 31.1% | 28 | 25.7% | 49 | 35.8% | 36 | 24.2% | 13 | 32.5% |
|  | Unknown | 21 | 8.4% | 12 | 11.0% | 8 | 5.8% | 12 | 8.1% | 4 | 10.0% |
| COVID-19 vaccination | Not received | 19 | 7.6% | 4 | 3.7% | 14 | 10.2% | 9 | 6.0% | 1 | 2.5% |
|  | Received | 232 | 92.4% | 105 | 96.3% | 123 | 89.8% | 140 | 94.0% | 39 | 97.5% |
|  | Unknown | 0 | 0.0% | 0 | 0.0% | 0 | 0.0% | 0 | 0.0% | 0 | 0.0% |
| CURB65†† | Low | 134 | 53.4% | 44 | 40.4% | 86 | 62.8% | 81 | 54.4% | 15 | 37.5% |
|  | Moderate | 87 | 34.7% | 46 | 42.2% | 40 | 29.2% | 52 | 34.9% | 16 | 40.0% |
|  | Severe | 30 | 12.0% | 19 | 17.4% | 11 | 8.0% | 16 | 10.7% | 9 | 22.5% |
| COPD | no | 171 | 68.1% | 68 | 62.4% | 100 | 73.0% | 69 | 46.3% | 25 | 62.5% |
|  | yes | 80 | 31.9% | 41 | 37.6% | 37 | 27.0% | 80 | 53.7% | 15 | 37.5% |
| Asthma | no | 184 | 73.3% | 89 | 81.7% | 93 | 67.9% | 83 | 55.7% | 33 | 82.5% |
|  | yes | 67 | 26.7% | 20 | 18.3% | 44 | 32.1% | 66 | 44.3% | 7 | 17.5% |
| Bronchiectasis | no | 250 | 99.6% | 109 | 100.0% | 137 | 100.0% | 148 | 99.3% | 40 | 100.0% |
|  | yes | 1 | 0.4% | 0 | 0.0% | 0 | 0.0% | 1 | 0.7% | 0 | 0.0% |
| IHD | no | 221 | 88.0% | 97 | 89.0% | 119 | 86.9% | 132 | 88.6% | 31 | 77.5% |
|  | yes | 30 | 12.0% | 12 | 11.0% | 18 | 13.1% | 17 | 11.4% | 9 | 22.5% |
| Hypertension* | no | 219 | 87.3% | 97 | 89.0% | 117 | 85.4% | 133 | 89.3% | 36 | 90.0% |
|  | yes | 32 | 12.7% | 12 | 11.0% | 20 | 14.6% | 16 | 10.7% | 4 | 10.0% |
| On immunosuppression | no | 199 | 79.3% | 85 | 78.0% | 111 | 81.0% | 107 | 71.8% | 36 | 90.0% |
|  | yes | 52 | 20.7% | 24 | 22.0% | 26 | 19.0% | 42 | 28.2% | 4 | 10.0% |
| Diabetes type | None | 199 | 79.3% | 88 | 80.7% | 107 | 78.1% | 116 | 77.9% | 28 | 70.0% |
|  | Type 1 | 3 | 1.2% | 2 | 1.8% | 0 | 0.0% | 1 | 0.7% | 0 | 0.0% |
|  | Type 2 | 49 | 19.5% | 19 | 17.4% | 30 | 21.9% | 32 | 21.5% | 12 | 30.0% |
| CKD** | None | 185 | 73.7% | 78 | 71.6% | 103 | 75.2% | 113 | 75.8% | 28 | 70.0% |
|  | Mild | 56 | 22.3% | 24 | 22.0% | 31 | 22.6% | 30 | 20.1% | 7 | 17.5% |
|  | Moderate and severe | 10 | 4.0% | 7 | 6.4% | 3 | 2.2% | 6 | 4.0% | 5 | 12.5% |

aLRTD, acute lower respiratory tract disease; CCI, Charlson comorbidity index; CKD, chronic kidney disease; COPD, chronic obstructive pulmonary disease; CRDE, chronic respiratory disease exacerbation; HF, heart failure; IHD, ischaemic heart disease; NP-LRTI, non-pneumonic lower respiratory tract infection; PCR, polymerase chain reaction; RSV, respiratory syncytial virus; SD, standard deviation
† In the UK, patients aged ≥65 years are eligible for Pneumococcal vaccination (PneumoVax®, PPV23) once, and annual influenza vaccine
\* Hypertension was only included if causing other cardiac complications
†† CURB-65 score 0-1= low, 2 = moderate and ≥3 = severe
\*\* Chronic kidney disease (CKD) was classified as mild if stage 1-3; moderate/severe if stage 4-5, end-stage renal failure or there was dialysis dependence

**Table 3:** Outcomes of adult patients hospitalised with aLTRD and test-positive for RSV. The median hospital length of stay, percentage intensive care or high-dependency unit (ICU/HDU) admission, median ICU/HDU length of stay, and percentage all-cause mortality within 30-days of admission among adults hospitalised with aLTRD and test-positive for RSV. These are reported as the percentage of patients test-positive for RSV among RSV tested patients.

| Variable | All included aLTRD | Tested for RSV | Test-positive for RSV |  |  |  |  |
| --- | --- | --- | --- | --- | --- | --- | --- |
|  | Total | Total | Total | Pneumonia | NP-LRTI | CRDE | HF |
| **Length of stay** |  |  |  |  |  |  |  |
| **Median days (IQR)** |  |  |  |  |  |  |  |
| Overall | 5 (2-13) | 6 (3-14) | 5 (2-8) | 6 (3-10) | 5 (2-7) | 5 (2-8) | 8 (4-16) |
| 18-64y | 3 (1- 7) | 4 (2- 9) | 4 (2- 7) | 4 (3-12) | 3 (2-6) | 4 (2- 6) | 6 (4-30) |
| 65-74y | 5 (2-12) | 6 (3-13) | 5 (3- 7) | 4 (2- 8) | 5 (3-6) | 4 (2- 6) | 10 (8-10) |
| ≥75y | 7 (3-18) | 8 (4-18) | 6 (3-10) | 6 (4-12) | 5 (2-8) | 6 (2-12) | 8 (3-12) |
| **ICU/HDU admission** |  |  |  |  |  |  |  |
| **N (%)** |  |  |  |  |  |  |  |
| Overall | 331/11445 (2.9%) | 228/6906 (3.3%) | 9/251 (3.6%) | 7/109 (6.4%) | 2/137 1.5%) | 4/149 (2.7%) | 3/40 (7.5%) |
| 18-64 | 150/3705 (4%) | 102/2224 (4.6%) | 4/76 (5.3%) | 3/28 (10.7%) | 1/45 (2.2%) | 1/43 (2.3%) | 1/11 (9.1%) |
| 65-74 | 92/2255 (4.1%) | 69/1533 (4.5%) | 0/48 (0%) | 0/21 (0%) | 0/27 (0%) | 0/38 (0%) | 0/3 (0%) |
| ≥75y | 89/5485 (1.6%) | 57/3149 (1.8%) | 5/127 (3.9%) | 4/60 (6.7%) | 1/65 (1.5%) | 3/68 (4.4%) | 2/26 (7.7%) |
| **ICU/HDU length of stay** |  |  |  |  |  |  |  |
| **Median days (IQR)** |  |  |  |  |  |  |  |
| Overall | 6 (3-10) | 6 (3-11) | 6 (3-13) | 6 (3-11) | 13 (8-14) | 10 (5-13) | 3 (2-4) |
| 18-64y | 6 (3-12) | 6 (3-12) | 13 (6-14) | 6 (4-18) | 14 (13-14) | 10 (8-11) | 6 (6- 6) |
| 65-74y | 6 (3-10) | 6 (3-11) | - | - | - | - | - |
| ≥75y | 6 (3-8) | 6 (4-8) | 4 (2- 7) | 5 (3- 9) | 3 (3- 3) | 7 (4-10) | 2 (2- 2) |
| **All-cause mortality** |  |  |  |  |  |  |  |
| **N (%)** |  |  |  |  |  |  |  |
| Overall | 1000/11445 (8.7%) | 494/6906 (7.2%) | 11/251 (4.4%) | 9/109 (8.3%) | 2/137 (1.5%) | 5/149 (3.4%) | 6/40 (15%) |
| 18-64y | 78/3705 (2.1%) | 43/2224 (1.9%) | 1/76 (1.3%) | 1/28 (3.6%) | 0/45 (0%) | 1/43 (2.3%) | 1/11 (9.1%) |
| 65-74y | 170/2255 (7.5%) | 93/1533 (6.1%) | 1/48 (2.1%) | 1/21 (4.8%) | 0/27 (0%) | 1/38 (2.6%) | 0/3 (0%) |
| ≥75y | 752/5485 (13.7%) | 358/3149 (11.4%) | 9/127 (7.1%) | 7/60 (11.7%) | 2/65 (3.1%) | 3/68 (4.4%) | 5/26 (19.2%) |

aLRTD, acute lower respiratory tract disease; CRDE; chronic respiratory disease exacerbation; HF, heart failure; ICU, intensive care unit; IQR, interquartile range; NP-LRTI, non-pneumonic lower respiratory tract infection; N, number; RSV, respiratory syncytial virus; y, years

The overall RSV test-positive-rate in hospitalised adults was 4.4% (95%CI:3.9-5.0%), based on SOC testing, and 3.6% (3.2-4.1) when research testing was added (Table 1). The RSV test-positive-rate trended towards increasing with patient age, and across all age groups we found more RSV test-positives in the NP-LRTI subgroup than in the pneumonia subgroup (Table 2). The RSV test-positivity rate showed a seasonal pattern, with modelled rates peaking in December between 7.9-12.7% (Fig. 3a). When stratified by age, the modelled test-positive seasonal peak was most clearly seen, and was latest, in the oldest age group (≥75y; Fig. 3b). During the off-peak season the SOC test-positivity was 1.4% (1.0-1.9), and the research lab test-positivity was 0.5% (0.2-0.9).

**Figure 3:**
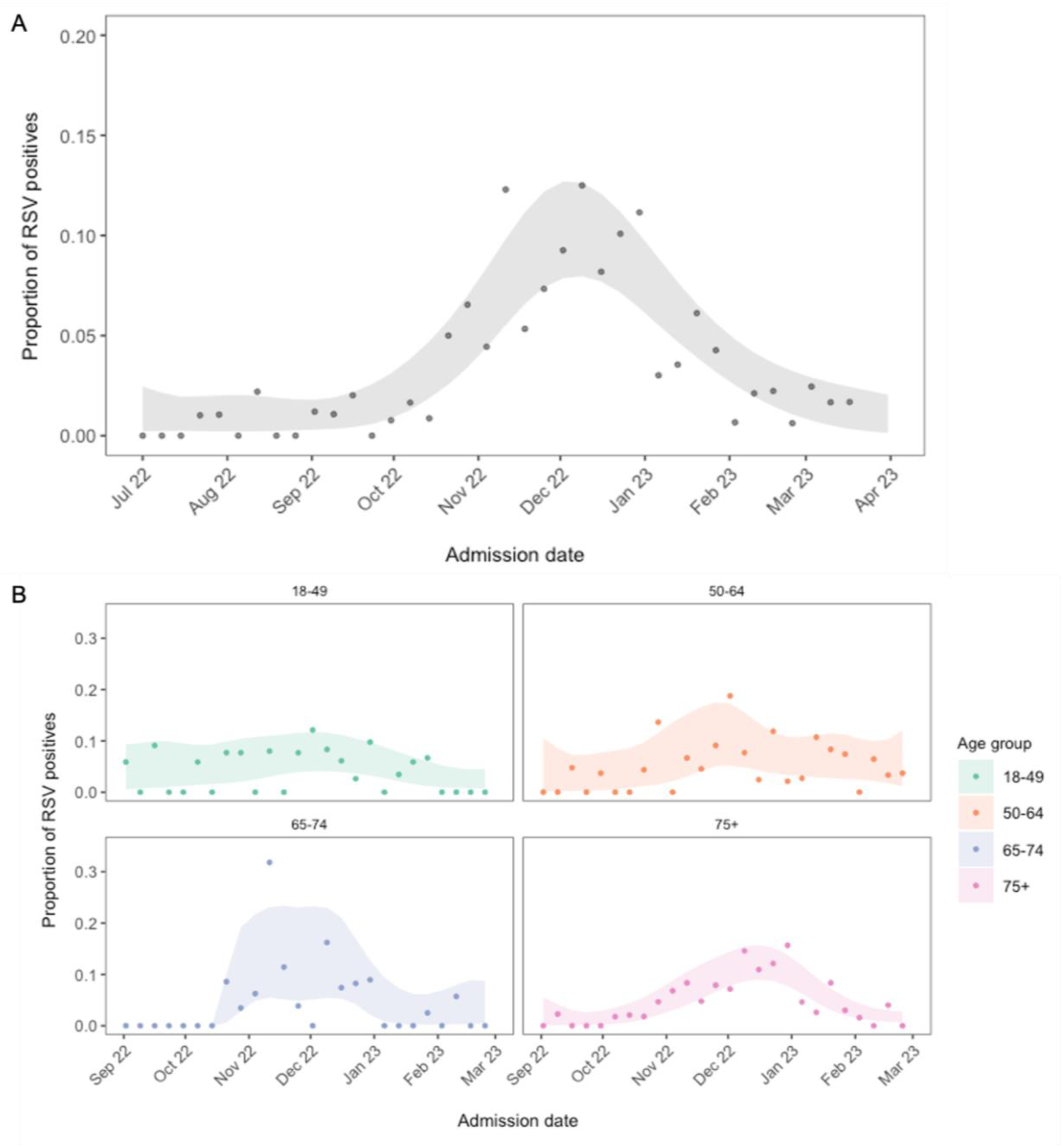
RSV test-positivity over time. The proportion of RSV test-positives over time, overall between July – April (A) and stratified by age group between September - March (B). Points show observed test-positive-rates for each week (participants RSV test-positive / participants RSV tested), and the ribbons show estimated uncertainty from a locally fitted quasi-binomial model.

### Adjustment for test error

The latent class (LC) model performed well on simulated data with known parameters, where test sensitivities and specificities were assumed to be the same (Supplementary, Data 1;2), when test sensitivities and specificities varied (Supplementary, Data 3), and when dependence was modelled between the research tests (Supplementary, Data 4). Model inferences were not sensitive to changes in the prior distribution for specificity (Supplementary, Data 5). Small differences between inferred and simulated sensitivity did not impact inferred prevalence.

The LC model inferred a prevalence of 2.3% (95%CI:1.4–3.7%) in the tested population, compared with an observed prevalence (test-positivity) of 3.6% (3.2-4.1; 251/6906). Test-positivity overestimated true prevalence in all age- and season-stratified subgroups (Fig. 4). The LC model for the peak RSV season (Nov-Feb) inferred a prevalence of 5.8% (3.5–9.2), compared to test-positivity of 7.9% (6.9–9.1). The model inferred a higher sensitivity and lower specificity for the SOC test and the sputum research lab sample compared to the NP/OP and saliva research lab samples (Table 4). These trends were common across subgroups, although uncertainty in test sensitivity was large (Extended Data, 6). The model inferred similar estimates of prevalence, sensitivity and specificity, overall, and for different participant subgroups, when SOC testing was split into the three different panel tests used, and when only the first SOC RSV test was included (Supplementary, Data 6;7).

**Figure 4:**
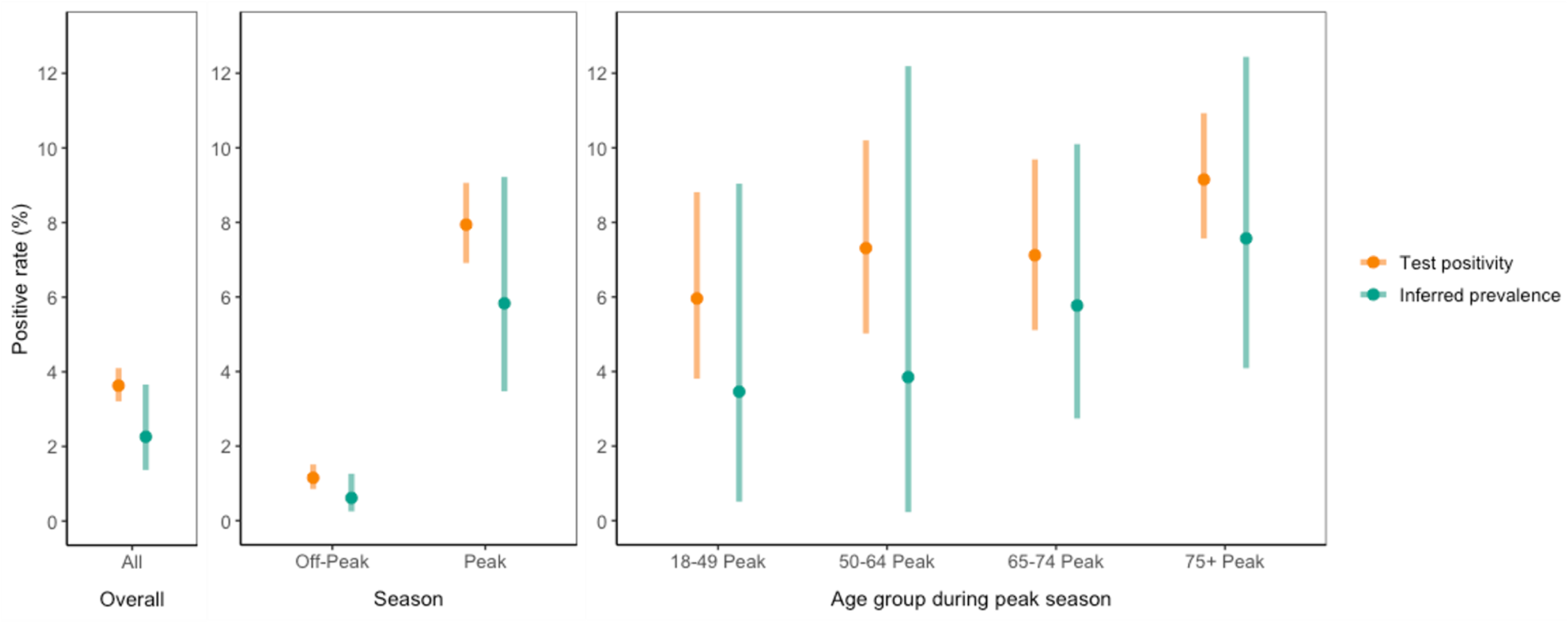
Test-positivity vs inferred true prevalence in different participant subgroups. Point estimates of test-positivity (any positive / tested; orange) and inferred true prevalence from a latent class model accounting for test error and multiple testing (mean; green), run for different participant subgroups split by peak season (Nov-Feb), off-peak season, and age groups during the peak season. Bars show 95% CI’s (orange: Clopper and Pearson confidence intervals; green: credible interval).

**Table 4:**
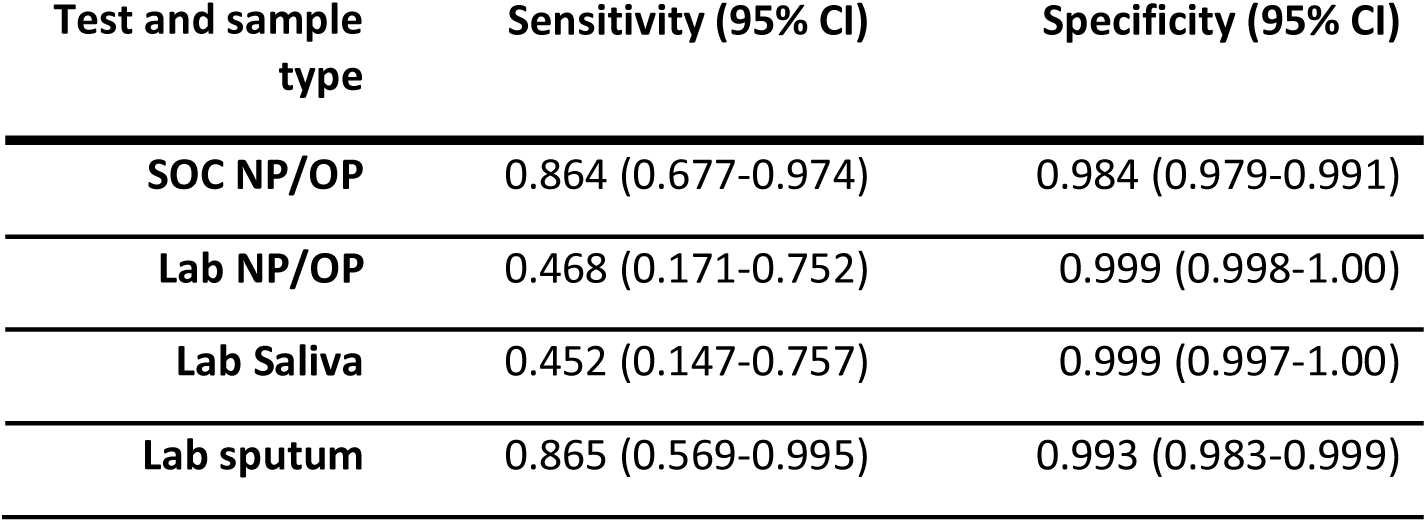
Latent class inferred test sensitivities and specificities. Sensitivity and specificity median inferences from study data (N= 6906) of 4 different tests and sample types for RSV (Standard of care (SOC NP/OP); and three research lab tests: (NP/OP; saliva, and sputum)) from a Bayesian latent class model (mean across chains). The model includes a dependence structure between the three research lab samples. 95% credible intervals are shown. The LC model inferred a prevalence of 2.3% (95%CI 1.4 – 3.7%).

At the low prevalence rates observed in this study, test sensitivity had little impact on accuracy of prevalence estimations, and overestimation of prevalence was driven by imperfect test specificity (Extended Data, 7; Fig. 5). A Rogan-Gladen estimator [47] demonstrated, at the inferred sensitivity and specificity of the SOC test, observed prevalence would not be an overestimation of true prevalence unless observed prevalence was >15% (Extended Data, 8). Therefore, combining highly specific research lab samples with SOC testing, resulted in a lower combined test specificity and PPV (Fig. 5).

**Figure 5:**
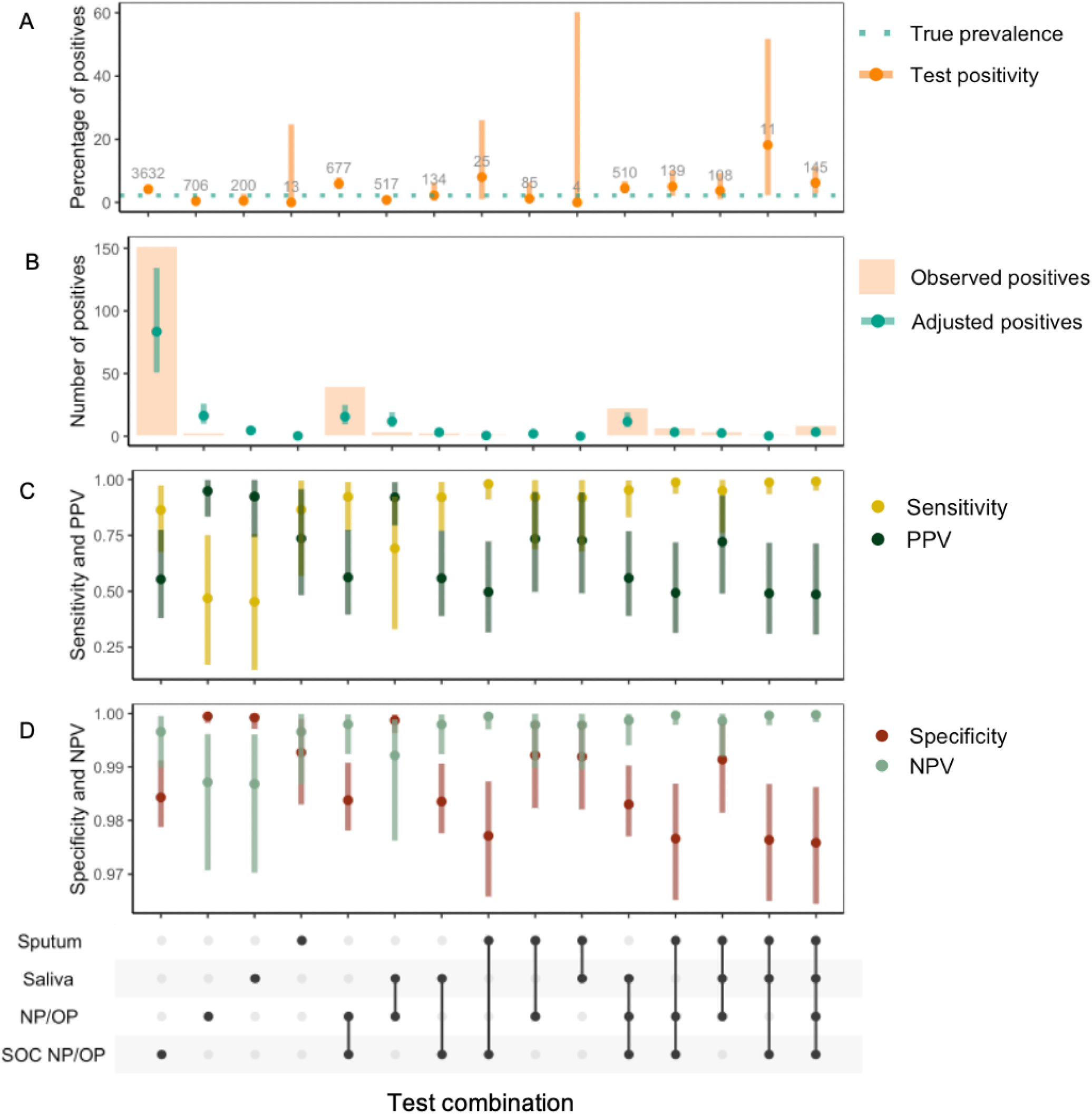
Test characteristics by different combinations of respiratory specimens. UpSet diagrams showing individuals tested by different mutually exclusive test/sample combinations of standard of care (SOC) naso- or oropharyngeal (NP/OP) respiratory swabs and research samples: sputum, saliva or NP/OP respiratory swabs. A) Percentage of RSV test-positive individuals (95%CI Clopper-Pearson). Numbers show number of participants tested. Dotted line shows true prevalence inferred from a latent class (LC) model; B) Number of RSV test-positive individuals. Points show number of RSV-positives after adjustment for true prevalence inferred from the LC model (with 95% credible intervals); C) Sensitivity and positive predictive values (PPV); D) Specificity and negative predictive values (NPV) from LC model (with 95% credible intervals).

### RSV incidence

After adjustment of the test-positivity for test error, and multiple testing, and accounting for the untested population, the overall calculated adult population RSV incidence/1000-person-years was 0.33 (95%CI:0.21–0.49). This ranged from 0.06 (0.0–0.12) in the youngest age group (18-49y), to 2.02 (1.10–3.06) in the oldest age group (≥75y; Table 5).

**Table 5:** Annual incidence of aLRTD hospitalised patients and RSV / 1000-person years. Incidence of aLRTD hospitalisation and hospitalised RSV per 1000-person-years (PYs), stratified by age group. RSV incidence is based on inferred prevalence of RSV from latent class models which adjust for test error and multiple testing. Latent class models were run separately for each age group for the peak RSV season, and an overall model was run for the off-peak season. The number of positives in each age group is the number of positives in the peak and off-peak season combined (see methods for details). 95% CIs are shown in brackets (95% credible intervals for the combined distribution of number of positives for each age group).

|  |  | All included aLRTD hospitalised |  |  |  |  | RSV hospitalised |
| --- | --- | --- | --- | --- | --- | --- | --- |
|  |  | Total (all-cause) | Pneumonia | NP-LRTI | CRDE | HF | Total |
| Age | Population | per 1000 PYs | per 1000 PYs | per 1000 PYs | per 1000 PYs | per 1000 PYs | per 1000 PYs |
| 18-49 | 435136 | 4.10 | 1.30 | 2.30 | 1.80 | 0.20 | 0.06 (0.02 - 0.12) |
| 50-64 | 157846 | 12.20 | 4.90 | 5.60 | 6.50 | 1.50 | 0.16 (0.03 - 0.41) |
| 65-74 | 72997 | 30.90 | 13.50 | 12.60 | 17.50 | 5.60 | 0.64 (0.34 - 1.04) |
| ≥75 | 70805 | 77.50 | 36.90 | 30.60 | 32.20 | 19.00 | 2.02 (1.10 - 3.06) |
| Overall | 736784 | 15.50 | 6.70 | 6.70 | 7.30 | 2.80 | 0.33 (0.21 - 0.49) |

## DISCUSSION

This prospective study found 251/6906 (3.6%; 95%CI:3.2-4.1%) of adults hospitalised with community-acquired aLRTD and tested for RSV in Bristol, UK, were test-positive. We modelled a weekly maximum test-positivity of 7.9-12.7%, with the seasonal peak more prominent in older adults. Recorded rates of RSV test-positivity generally increased with patient age, and a higher RSV test-positive-rate was found among NP-LRTI than pneumonia cases. Over the 12-month study period, 4.2% of CRDE cases and 3.2% of HF cases, that were tested for RSV, were RSV test-positive.

The epidemiology of RSV was disrupted by social distancing measures implemented to reduce infections and hospitalisations following SARS-CoV-2 emergence, with subsequent out-of-season peaks, probably caused by waning population immunity [48–50]. Our analysis is the first UK study to report the RSV-positive-rate following SARS-CoV-2 emergence, and in adults hospitalised with ARI across aLRTD and its subgroups. We show that between April 2022-March 2023, the RSV test-positive peak in adults had returned to the standard winter season, peaking between Nov–Feb, consistent with trends observed across 2022/23 by UKHSA sentinel surveillance [50]. The maximum modelled weekly RSV test-positivity of 7.9-12.7% in all adults is comparable to the 13.7% maximum reported SARS-CoV-2 positive-rate in 65-year-olds in 2022-23 in the UK, but lower than the 26.1% recorded for influenza in ≥65-year-olds [50]. Our incidence estimates’ confidence intervals (0.34-1.04 [65-74y]; 1.10-3.06 [≥75y]/1000 population) encompass other modelled estimates for the UK, with estimated RSV hospitalisation incidence/1000 population of 0.77 (0.46-1.09) [21] in 65-74-year-olds, and between 1.16 (0.91-1.48) and 3.38 (2.50-4.05) for low and high risk adult populations, respectively, in >75-year-olds (with risk populations based on chronic conditions indicative of severe influenza risk)[13]. Our incidence estimates fall within these low and high risk adult population estimates for all age groups [13]. Our estimates are comparable to other European countries, with overall modelled hospitalisation incidence/1000 population of 0.66 (0.55-0.76) in 65-75-year-olds [21], and to other high-income countries worldwide, with a pooled meta-analysis estimating 0.9-2.2 hospitalisation incidence/1000 population in >60-year-olds [51]. This study adds to the small number of prospective studies quantifying adult RSV burden outside of the US [52].

The LCA performed well in inferring both prevalence and test error parameters. Prevalence estimates are overwhelmingly affected by test specificity in our study. Test sensitivity had very little impact on observed prevalence. At the parameter ranges in this study, observed prevalence always overestimated true prevalence (Extended Data, 7). This is expected when the disease of interest is uncommon, as underlying false positive-rates, even if they are low, have a high relative effect on the true positive-rate [22]. This effect will be accentuated when multiple tests are done if all positive results are assumed to be true positives, as this increases overall sensitivity at the cost of overall specificity [25], resulting in a lower positive predictive value of the combination of multiple tests than the individual tests. The PPV will be particularly low in studies, such as this one, that include out-of-season testing for RSV when infection rates are especially low [47]. In our study, most RSV-positive individuals were detected by SOC testing (232/251), therefore it is expected that imperfect SOC specificity (inferred to be 98.4%), will result in a significant proportion of test-positives being false positives (Fig. 5). Thus, even in the absence of RSV in the population, we would expect to see a test-positivity rate of 1.6% representing test noise. Consistent with this, in our off-season population, the SOC positivity-rate was 1.4%. As RSV incidence in adults is generally low [21;51;52], adjustment of test-positivity for both the false negative and false positive-rate, as demonstrated here, is important for accurate estimates of adult RSV burden in studies worldwide.

Factors other than test error will also affect accurate ascertainment of true population prevalence. Studies only investigating pneumonia will exclude cases found in NP-LRTI, HF and CRDE, and we note the RSV positivity-rate was higher in NP-LRTI than pneumonia-LRTI and confirm that RSV is associated with CRDE and HF. Comorbid disease, and hence the risk posed by RSV infection [53] accumulates with patient age; however, older patients are less likely to undergo SOC testing [17]. This may disproportionately under-estimate RSV burden among the patient groups most at risk. In this study, research samples were taken an average of 2-days later than SOC samples (Extended Data, 5), so RSV titres in research samples would be predicted to be lower, contributing to the low inferred sensitivities of these tests. However, the LC approach infers sensitivities/specificities of the end-to-end testing system (as opposed to laboratory test sensitivities/specificities), accounting for differences in pre-test probability caused by different participant selection criteria for testing, sampling errors, and the time between sampling and laboratory handling.

The lower sensitivity of the research test in our study means adding sputum or saliva sample testing to NP/OP samples increased apparent RSV detection. This aligns with previous studies that found adding both saliva and sputum sample results increased apparent RSV detection by 63% over NP results alone [19], while adding sputum or saliva test results alone increased apparent detection by 39-52% [19;18;54] and 55% [19], respectively, assuming perfect test specificity [19]. Of practical relevance, all these specimens may be obtained during acute illness and thus results may inform patient management, whereas serology requires a convalescent sample. Saliva can be a reliable and sensitive specimen type for RSV and other respiratory viruses [19;55]. Our research lab NP/OP and saliva samples both had perfect specificity, so combining these results would increase true RSV detection (Fig. 5). We found the sensitivity of sputum samples was ∼twice as high as for NP/OP swabs. In older adults, sputum has stronger RSV [56–57], influenza [58], and coronavirus [58–59] PCR signals than nasal swabs. Taken together, these results suggest that future studies relying solely on low sensitivity and high specificity tests, could achieve more accurate estimates of RSV disease burden by testing multiple specimen types. However, if any test has imperfect specificity, such multiple testing will amplify this and result in overestimation of true RSV prevalence, unless adjusted for in the way we demonstrate here.

Our study has many strengths, including the comprehensive ascertainment of LRTI in adults hospitalised within a defined geographical area. Notably, we undertook case-by-case ascertainment, the epidemiological gold-standard [60], and did not rely on clinical coding, a microbiological database or modelling alone to estimate RSV disease. Inclusion of patient data obtained under Section 251 of the 2006 NHS Act allows us to make accurate comparisons and draw appropriate conclusions about the representativeness of our sample. We enrolled adults who lacked capacity through a consultee, undertaking enhanced research sampling and avoiding under-representing these patients in this study, including those with aLRTD who are severely ill or have advanced dementia or other frailty.

The study also has some limitations. Despite undertaking research testing to increase the proportion tested, 40% of the cohort were not tested for RSV. However, the proportion of untested participants was lower (26%) during the peak season (Nov-Feb), in part due to increased clinical suspicion resulting in increased SOC testing during this period. We found statistically significant differences between RSV tested and untested patients, and therefore the underlying prevalence in these two groups is likely to differ due to differing patient characteristics. For population incidence calculations, inferred prevalence from the LC models were assumed to be the same in the equivalent untested population subgroup. Stratifying by age group and season controls for the biggest factors affecting clinical suspicion and pre-test probability, especially as RSV is characterised by non-specific respiratory symptoms. Nevertheless, without data on the rationale behind testing decisions, more precise differences between the underlying prevalence in the tested and untested population cannot be adjusted for. Finally, this study was a single-centre study, and while the Bristol population is representative of the UK, it is predominantly White-British (84%;[61]) and therefore aLRTD, RSV disease and RSV testing practices in other cohorts may vary from those reported here.

In conclusion, this prospective study is the first to estimate the RSV burden among adults hospitalised with aLRTD and its subgroups in the UK post-COVID. We found that 3.6% (95%CI:3.2-4.1) of annual RSV tested aLRTD hospital cases were RSV-positive, with modelled weekly test-positivity peaking at 7.9-12.6%. Test-positivity rates increased with patient age, were associated with a 4.4% all-cause 30-day mortality among hospitalised adults, and 3.6% of adults hospitalised with an RSV-positive test required ICU-level care. Test-positivity (3.6%) overestimated true prevalence (2.3%), due to the low positivity-rates and imperfect test specificity, which was exacerbated by out-of-season and multiple testing. After LC adjustment for test characteristics, we estimated overall adult population incidence/1000-person-years to be 0.33 (0.21-0.49), and 2.02 (1.10-3.06) in ≥75-year-olds. Therefore, implementation of effective RSV vaccines has the potential to reduce hospitalised adult aLTRD morbidity and mortality, especially among the elderly, and the accuracy of future studies of adult RSV burden and vaccine-effectiveness can be improved by adjustment for error introduced by test characteristics.

### Data Sharing

The data used in this study are sensitive and cannot be made publicly available without breaching patient confidentiality rules. Therefore, individual participant data and a data dictionary are not available to other researchers.

### Ethics and permission

This prospective observational cohort study of adults admitted to two large university hospitals (Southmead, and Bristol Royal Infirmary) in Bristol, UK, was approved by the Health Research Authority reference 20/EE/0157, ISRCTN: 17354061. Informed consent was obtained from patients with capacity, and declarations for participation from consultees for individuals lacking capacity. If it was not practicable to approach individuals for consent, data were included using approval from the Clinical Advisory Group under Section 251 of the 2006 NHS Act.

## Contributors

KL, RC, AC, GQ, AF, LD and CH generated the research questions and analysis plans. The AvonCAP research team, CH, JK, BM and SM were involved in data collection. CH, JK, SM and AF verified the data. KL, RC, AC, GO, GQ, AF, LD and CH undertook data analysis. All authors (KL, RC, AC, GQ, GO, JK, SM, BM, KD, ARP, WH, JO, NM, AF, LD, and CH) contributed to preparation of the manuscript and its revisions before publication. AF and CH provided oversight of the research.

## Acknowledgements

We thank colleagues at the University of Bristol for their support with this study, including Rachel Davies, Paul Savage, Emma Foose, Susan Christie, Mark Mummé, Alison Horne, Mai Baquedano, and Adam Taylor. We would like to thank Stewart Robinson, David Clint and Henry Stuart and their teams for the support provided during this study. We would also like to acknowledge the research teams at North Bristol and University Hospitals of Bristol and Weston NHS Trusts for making this study possible, including Helen Lewis-White, Rebecca Smith, Rajeka Lazarus, Jane Blazeby, Diana Benton, and David Wynick. Finally, we would like to thank Aman Kaur-Singh and Kevin Sweetland for their support with this study.

## The AvonCAP Research Group

Aaran Sinclair, Ainhoa Rodriguez-Pereira, Amelia Langdon, Amy Taylor, Anabella Turner, Anna Jones, Anna Koi, Anya Mattocks, Begonia Morales-Aza, Bethany Osborne, Brianna Dooley, Callum Hawkins, Charli Grimes, Chloe Farren, Christian Povey, Claire Mitchell, David Adegbite, Dylan Thomas, Elinor Balch, Ella Ackroyd-Weldon, Emma Bridgeman, Emma Scott, Felix Wright, Ffion Davies, Fiona Perkins, Francesca Bayley, Gabriella Ruffino, Gabriella Valentine, Georgina Mortimer, Grace Tilzey, Harriet Ibbotson, Hanah Batholomew, Hugo Swift, Jacob Symanowski, Ilana Kelland, Imogen Ely, Jade King, Jake Whittle, Jane Kinney, Johanna Kellett Wright, Jonathan Vowles, Josephine Bonnici, Josh Anderson, Juan Garcia-Tello, Julia Brzezinska, Julie Cloake, Kaltun Duale, Katarina Milutinovic, Kate Helliker, Katie Maughan, Kazminder Fox, Kellie Pettinger, Konstantina Minou, Kyla Chandler, Lana Ward, Leah Fleming, Leigh Morrison, Liberty Smith, Lily Smart, Lisa Grimmer, Louise Setter, Louise Wright, Lucy Grimwood, Maddalena Bellavia, Madeleine Clout, Maia Lyall, Malak Eghleilib, Marianne Vasquez, Maria Garcia Gonzalez, Mariella Ardeshir, Marta Mergulhao, Martina Chmelarova, Matthew Randell, Michael Booth, Milo Jeenes-Flanagan, Miriama Resutikova, Monika Chaulagain, Natalie Chang, Nefeli Tavira, Nellie Farhoudi, Niall Grace, Nicola Manning, Oliver Griffiths, Olivia Pearce, Pip Croxford, Peter Sequenza, Petronela Anchidin, Rajeka Lazarus, Rebecca Clemence, Rosa Aldridge, Rhian Walters, Riley Cooper, Robin Marlow, Robyn Heath, Rupert Antico, Sandi Nammuni Arachchge, Sarah Stollery, Seevakumar Suppiah, Sean Robinson, Serena McGuinness, Siddiqa Uddin, Taslima Mona, Tawassal Riaz, Teagan Barrett, Tom Long, Tudor Dimofte, William Healy, Yassin Ben Khoud, Vicki Mackay, Zahra Hashmi, Zandile Maseko, Zoe Taylor, Zsuzsa Szasz-Benczur, Zsolt Friedrich.

## Extended Data

**Extended data 1:** Case definitions. aLRTD included infectious and non-infectious clinical diagnoses and was defined as any presentation with acute lower respiratory disease, including pneumonia; NP-LRTI; acute bronchitis; exacerbations of underlying cardiorespiratory disease (CRDE) including chronic obstructive pulmonary disease (COPD) and asthma; and acute or decompensated heart failure (HF). aLRTD patients were designated RSV test-positive if they tested positive on PCR, from any specimen type. Pneumonia was defined as acute respiratory illness with radiological changes compatible with infection or when the treating clinician confirmed this diagnosis or both, in line with NICE/BTS guidelines [39–40]. NP-LRTI was defined as the presence of signs and symptoms of acute lower respiratory tract infection in the absence of both indicative radiological infiltrates and a clinical pneumonia diagnosis. Under these case definitions, any patients with aLRTD signs or symptoms due to non-infectious aLRTD were assigned appropriately to CRDE or HF groups.

| Condition | Definition | Reference |
| --- | --- | --- |
| Acute Lower Respiratory Tract Disease (aLRTD) | Acute lower respiratory tract disease (aLRTD) encompasses pneumonia, lower respiratory tract infection (LRTI), acute bronchitis, exacerbation of underlying respiratory disease including asthma and chronic obstructive pulmonary disease (COPD). Pneumothorax, pulmonary embolism, progression or new diagnosis of primary or secondary lung malignancy were excluded from aLRTD. | Hyams *et al*, BMJ Open 2022; 12:e057464. doi: 10.1136/bmjopen-2021-057464BMJ Open paper |
| Pneumonia | Pneumonia was defined as infection affecting the airways (below the level of the larynx), with either (1) an acute illness with radiographic shadowing which was at least segmental or present in more than one lobe and was not known to be previously present or due to other causes, OR (2) in the absence of radiological investigation, clinical confirmation of pneumonic disease in the opinion of the treating physician | Lim WS, Baudouin SV, George RC, *et al*; Pneumonia guidelines Committee of the BTS Standards of Care Committee. Thorax. 2009 Oct;64 Suppl 3:iii1-55. |
| Non-Radiologically Proven Lower Respiratory Tract Infection (NP-LRTI) | An infection that affects the airways (below the level of the larynx) including the trachea and alveoli, with neither the presence of radiological change nor a clinical diagnosis of pneumonia from the treating physician, i.e. non-pneumonic infection in the lungs. | Anderson W, Winter J. Managing LRTI in adults in the community. Practitioner. 2009 Nov;253(1723):21-5, 2-3. PMID: 20043506. |
| Cardiac/Heart Failure (HF) | A clinical syndrome with symptoms and/or signs caused by a structural and/or functional cardiac abnormality and corroborated by elevated natriuretic peptide levels and or objective evidence of pulmonary or systemic congestion. | Bozkurt, Biykem *et al*. Universal Definition and Classification of Heart Failure. Journal of Cardiac Failure, Volume 27, Issue 4, 387 – 413. |
| Chronic Respiratory Disease Exacerbation (CRDE) | An acute ($\leq 28$ day) deterioration in respiratory function with the presence of signs/symptoms of aLRTD in a patient with a chronic respiratory disease, in line with the accepted definition of an acute exacerbation for the given chronic respiratory definition. | |

**Extended data 2:**
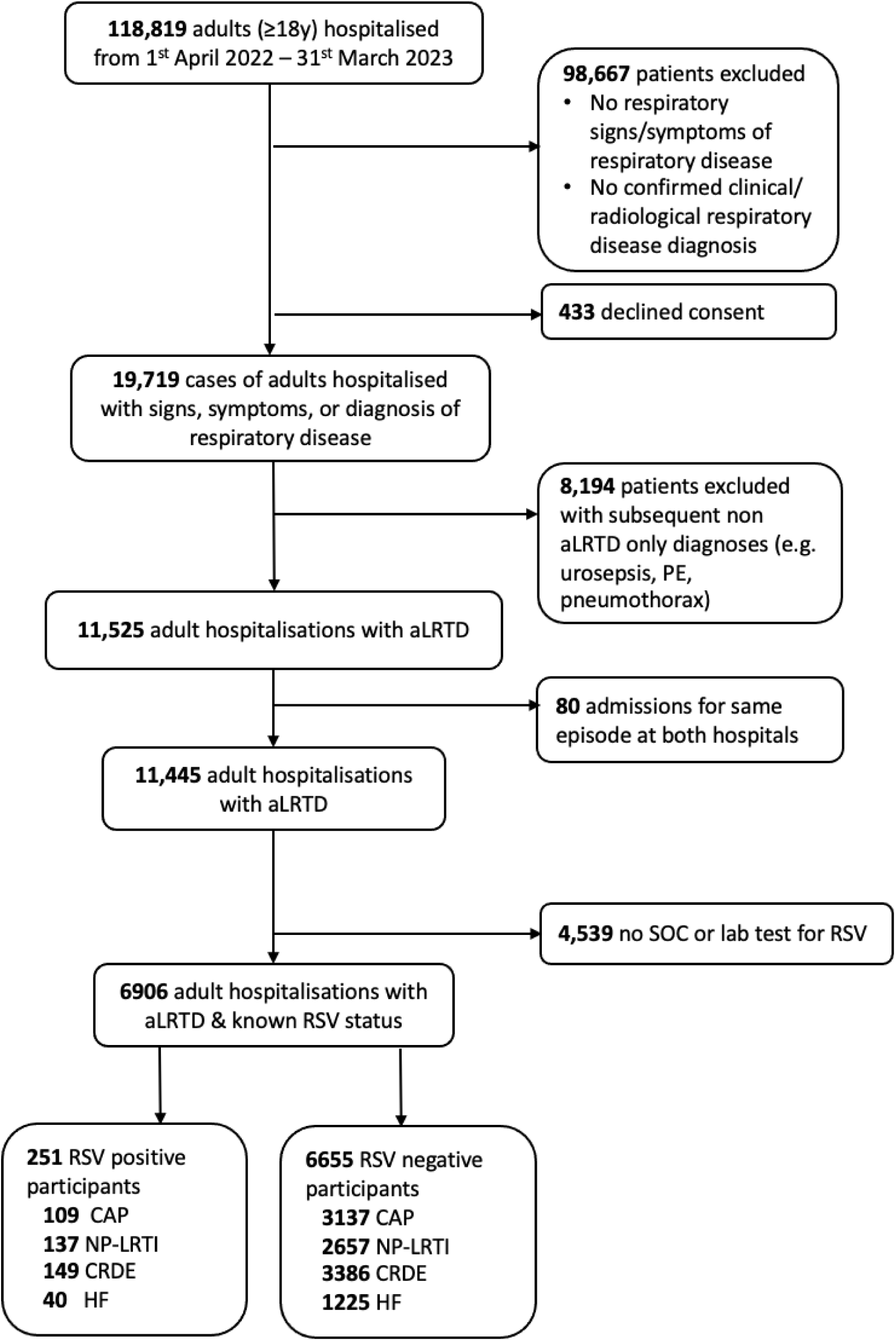
Study flow diagram. aLRTD, acute lower respiratory tract disease; CAP, community-acquired pneumonia; CRDE, chronic respiratory disease exacerbation; HF, heart failure; NP-LRTI, non-pneumonic lower respiratory tract infection; PE, pulmonary embolus; RSV, Respiratory syncytial virus; SOC, standard-of-care. CAP, NP-LRTI, CRDE, and HF are not mutually exclusive groups.

**Extended data 3:** Patient characteristics of adults hospitalised with aLRTD, tested for and positive for RSV by standard of care (SOC) or research lab. Patient demographics are shown for all adults hospitalised with aLRTD during the study period, patients tested for RSV infection by any test (SOC NP/OP or Research lab NP/OP, saliva, or sputum samples), patients not tested for RSV, and patients positive for RSV. Full demographics by RSV testing are presented in Extended data 4. The differences in patient characteristics for each group between those tested for RSV and not, were tested by Kolmogorov–Smirnov or Fisher’s tests and P values are given.

|  |  | All included aLTRD |  | Tested for RSV<br>(SOC or Research lab) |  | Not tested for RSV |  | P value |
| --- | --- | --- | --- | --- | --- | --- | --- | --- |
| Variable | Characteristic | Total |  | Total |  | Total |  |  |
|  |  | N | Value | N | Value | N | Value |  |
| Age | Median (IQR) | 11445 | 74.2<br>(59.8-83.8) | 6906 | 73.4 (60.3-82.5) | 4539 | 75.6 (59.0-85.7) | — |
| Age category (y) † | 18-34 | 781 | 6.8% | 411 | 6.0% | 370 | 8.2% | <0.001 |
|  | 35-49 | 1000 | 8.7% | 603 | 8.7% | 397 | 8.7% |  |
|  | 50-64 | 1924 | 16.8% | 1210 | 17.5% | 714 | 15.7% |  |
|  | 65-74 | 2255 | 19.7% | 1533 | 22.2% | 722 | 15.9% |  |
|  | 75-84 | 2943 | 25.7% | 1830 | 26.5% | 1113 | 24.5% |  |
|  | ≥85 | 2542 | 22.2% | 1319 | 19.1% | 1223 | 26.9% |  |
| Gender | Male | 5538 | 48.4% | 3358 | 48.6% | 2180 | 48.0% | 0.54 |
|  | Female | 5907 | 51.6% | 3548 | 51.4% | 2359 | 52.0% |  |
| Pneumonia | yes | 4942 | 43.2% | 3246 | 47.0% | 1696 | 37.4% | <0.001 |
| NP-LRTI | yes | 5390 | 47.1% | 2794 | 40.5% | 2159 | 47.6% | <0.001 |
| CRDE | yes | 4953 | 43.3% | 3535 | 51.2% | 1855 | 40.9% | <0.001 |
| HF | yes | 2063 | 18.0% | 1265 | 18.3% | 798 | 17.6% | 0.32 |
| Ethnicity | Asian | 194 | 1.7% | 111 | 1.6% | 83 | 1.8% |  |
|  | Black | 203 | 1.8% | 109 | 1.6% | 94 | 2.1% |  |
|  | Mixed origin | 138 | 1.2% | 89 | 1.3% | 49 | 1.1% |  |
|  | Other ethnicity | 93 | 0.8% | 46 | 0.7% | 47 | 1.0% |  |
|  | Unknown | 2058 | 18.0% | 1107 | 16.0% | 949 | 20.9% | <0.001 |
|  | White British | 8467 | 74.0% | 5270 | 76.3% | 3197 | 70.4% |  |
|  | White other | 292 | 2.6% | 172 | 2.5% | 120 | 2.6% |  |
| Care home resident | no | 10486 | 91.6% | 6409 | 92.8% | 4077 | 89.8% | <0.001 |
|  | yes | 959 | 8.4% | 497 | 7.2% | 462 | 10.2% |  |
| Smoker | Current smoker | 1527 | 13.3% | 980 | 14.2% | 547 | 12.1% | <0.001 |
|  | Ex-smoker | 5234 | 45.7% | 3254 | 47.1% | 1980 | 43.6% |  |
|  | Non-smoker | 3855 | 33.7% | 2240 | 32.4% | 1615 | 35.6% |  |
|  | Unknown | 829 | 7.2% | 432 | 6.3% | 397 | 8.7% |  |
| COVID-19 vaccination | Not received | 1323 | 11.6% | 733 | 10.6% | 590 | 13.0% | <0.001 |
|  | Received | 10074 | 88.0% | 6148 | 89.0% | 3926 | 86.5% |  |
|  | Unknown | 48 | 0.4% | 25 | 0.4% | 23 | 0.5% |  |
| CURB-65 | Low | 5871 | 51.3% | 3585 | 51.9% | 2286 | 50.4% | 0.0087 |
|  | Moderate | 3594 | 31.4% | 2187 | 31.7% | 1407 | 31.0% |  |
|  | Severe | 1980 | 17.3% | 1134 | 16.4% | 846 | 18.6% |  |
| CCI | Median (IQR) | 11445 | 4.0 (3.0-6.0) | 6906 | 4.0 (3.0-6.0) | 4539 | 4.0 (2.0-6.0) | 0.3 |
| COPD | no | 8035 | 70.2% | 4656 | 67.4% | 3379 | 74.4% | <0.001 |
|  | yes | 3410 | 29.8% | 2250 | 32.6% | 1160 | 25.6% |  |
| Asthma | no | 9596 | 83.8% | 5737 | 83.1% | 3859 | 85.0% | 0.0061 |
|  | yes | 1849 | 16.2% | 1169 | 16.9% | 680 | 15.0% |  |
| Bronchiectasis | no | 11400 | 99.6% | 6871 | 99.5% | 4529 | 99.8% | 0.02 |
|  | yes | 45 | 0.4% | 35 | 0.5% | 10 | 0.2% |  |
| IHD | no | 9945 | 86.9% | 5984 | 86.6% | 3961 | 87.3% | 0.35 |
|  | yes | 1500 | 13.1% | 922 | 13.4% | 578 | 12.7% |  |
| Hypertension* | no | 9849 | 86.1% | 5866 | 84.9% | 3983 | 87.8% | <0.001 |
|  | yes | 1596 | 13.9% | 1040 | 15.1% | 556 | 12.2% |  |
| On immunosuppression | no | 9969 | 87.1% | 5913 | 85.6% | 4056 | 89.4% | <0.001 |
|  | yes | 1476 | 12.9% | 993 | 14.4% | 483 | 10.6% |  |
| Diabetes type | None | 9131 | 79.8% | 5435 | 78.7% | 3696 | 81.4% | 0.0011 |
|  | Type 1 | 150 | 1.3% | 90 | 1.3% | 60 | 1.3% |  |
|  | Type 2 | 2164 | 18.9% | 1381 | 20.0% | 783 | 17.3% |  |
| CKD** | None | 8404 | 73.4% | 5111 | 74.0% | 3293 | 72.5% | 0.078 |
|  | Mild | 2587 | 22.6% | 1541 | 22.3% | 1046 | 23.0% |  |
|  | Moderate or Severe | 454 | 4.0% | 254 | 3.7% | 200 | 4.4% |  |

aLRTD, acute lower respiratory tract disease; CCI, Charlson comorbidity index; CKD, chronic kidney disease; COPD, chronic obstructive pulmonary disease; CRDE, chronic respiratory disease exacerbation; HF, heart failure; IHD, ischaemic heart disease; NP-LRTI, non-pneumonic lower respiratory tract infection; PCR, polymerase chain reaction; RSV, respiratory syncytial virus; SD, standard deviation
† In the UK, patients aged ≥65 years are eligible for Pneumococcal vaccination (PneumoVax®, PPV23) once, and annual influenza vaccine
\* Hypertension was only included if causing other cardiac complications
†† CURB-65 score 0-1= low, 2 = moderate and ≥3 = severe
\*\* Chronic kidney disease (CKD) was classified as mild if stage 1-3; moderate/severe if stage 4-5, end-stage renal failure or there was dialysis dependence

**Extended data 4:** Comparison of adults hospitalised with aLRTD by RSV testing. Patient demographics are shown for patients tested for RSV infection by both standard of care (SOC) NP/OP samples, and research lab samples (NP/OP, saliva, or sputum); just by SOC (SOC-only); and just by research lab (research-only). The differences in patient characteristics for each group between those tested by SOC-only, research lab-only, and both SOC and research lab were tested by Kolmogorov–Smirnov or Fisher’s tests and P values are given.

| Variable | Characteristic | Tested for RSV<br>(SOC and Research) |  | Tested for RSV<br>(SOC-only) |  | Tested for RSV<br>(Research-only) |  | Difference between SOC and<br>Research tested |
| --- | --- | --- | --- | --- | --- | --- | --- | --- |
|  |  | N | Value | N | Value | N | Value | P value |
| Age | Median (IQR) | 1641 | 70.8 (58.5-79.1) | 3632 | 74.6 (60.6-84.3) | 1633 | 73.5 (61.5-81.9) | <0.001 |
| Age category | 18-34 | 95 | 5.8% | 232 | 6.4% | 84 | 5.1% | <0.001 |
|  | 35-49 | 171 | 10.4% | 320 | 8.8% | 112 | 6.9% |  |
|  | 50-64 | 323 | 19.7% | 578 | 15.9% | 309 | 18.9% |  |
|  | 65-74 | 430 | 26.2% | 723 | 19.9% | 380 | 23.3% |  |
|  | 75-84 | 417 | 25.4% | 939 | 25.9% | 474 | 29.0% |  |
|  | 85+ | 205 | 12.5% | 840 | 23.1% | 274 | 16.8% |  |
| Gender | Male | 812 | 49.5% | 1690 | 46.5% | 856 | 52.4% | <0.001 |
|  | Female | 829 | 50.5% | 1942 | 53.5% | 777 | 47.6% |  |
| Pneumonia | yes | 802 | 48.9% | 1816 | 50.0% | 628 | 38.5% | <0.001 |
| CRDE | yes | 945 | 57.6% | 1763 | 48.5% | 827 | 50.6% | <0.001 |
| NPLRTI | yes | 638 | 38.9% | 1504 | 41.4% | 652 | 39.9% | 0.2 |
| HF | yes | 292 | 17.8% | 596 | 16.4% | 377 | 23.1% | <0.001 |
| Ethnicity | Asian | 18 | 1.1% | 76 | 2.1% | 17 | 1.0% | <0.001 |
|  | Black | 14 | 0.9% | 74 | 2.0% | 21 | 1.3% |  |
|  | Mixed origin | 19 | 1.2% | 55 | 1.5% | 15 | 0.9% |  |
|  | Other ethnicity | 14 | 0.9% | 26 | 0.7% | 6 | 0.4% |  |
|  | Unknown | 280 | 17.1% | 633 | 17.4% | 194 | 11.9% |  |
|  | White British | 1260 | 76.8% | 2661 | 73.3% | 1349 | 82.6% |  |
|  | White other | 35 | 2.1% | 106 | 2.9% | 31 | 1.9% |  |
|  | <missing> | 1 | 0.1% | 1 | 0.0% | 0 | 0.0% |  |
| Care home resident | no | 1579 | 96.2% | 3253 | 89.6% | 1577 | 96.6% | <0.001 |
|  | yes | 62 | 3.8% | 379 | 10.4% | 56 | 3.4% |  |
| Smoker | Current smoker | 246 | 15.0% | 491 | 13.5% | 243 | 14.9% | 0.021 |
|  | Ex-smoker | 797 | 48.6% | 1665 | 45.8% | 792 | 48.5% |  |
|  | Non-smoker | 500 | 30.5% | 1227 | 33.8% | 513 | 31.4% |  |
|  | Unknown | 98 | 6.0% | 249 | 6.9% | 85 | 5.2% |  |
| Covid vaccination | Not received | 159 | 9.7% | 384 | 10.6% | 190 | 11.6% | 0.15 |
|  | Received | 1478 | 90.1% | 3230 | 88.9% | 1440 | 88.2% |  |
|  | Unknown | 4 | 0.2% | 18 | 0.5% | 3 | 0.2% |  |
| CURB65 | Low | 960 | 58.5% | 1735 | 47.8% | 890 | 54.5% | <0.001 |
|  | Moderate | 449 | 27.4% | 1215 | 33.5% | 523 | 32.0% |  |
|  | Severe | 232 | 14.1% | 682 | 18.8% | 220 | 13.5% |  |
| CCI | Median (IQR) | 1641 | 4.0 (2.0-5.0) | 3632 | 4.0 (3.0-6.0) | 1633 | 4.0 (3.0-6.0) | <0.001 |
| COPD | no | 1060 | 64.6% | 2514 | 69.2% | 1082 | 66.3% | 0.0022 |
|  | yes | 581 | 35.4% | 1118 | 30.8% | 551 | 33.7% |  |
| Asthma | no | 1321 | 80.5% | 3036 | 83.6% | 1380 | 84.5% | 0.0047 |
|  | yes | 320 | 19.5% | 596 | 16.4% | 253 | 15.5% |  |
| Bronchiectasis | no | 1632 | 99.5% | 3622 | 99.7% | 1617 | 99.0% | 0.0048 |
|  | yes | 9 | 0.5% | 10 | 0.3% | 16 | 1.0% |  |
| IHD | no | 1413 | 86.1% | 3182 | 87.6% | 1389 | 85.1% | 0.032 |
|  | yes | 228 | 13.9% | 450 | 12.4% | 244 | 14.9% |  |
| Hypertension* | no | 1434 | 87.4% | 3047 | 83.9% | 1385 | 84.8% | 0.004 |
|  | yes | 207 | 12.6% | 585 | 16.1% | 248 | 15.2% |  |
| On immunosuppression | no | 1579 | 96.2% | 3253 | 89.6% | 1577 | 96.6% | <0.001 |
|  | yes | 62 | 3.8% | 379 | 10.4% | 56 | 3.4% |  |
| Diabetes type | None | 1291 | 78.7% | 2867 | 78.9% | 1277 | 78.2% | 0.19 |
|  | Type 1 | 27 | 1.6% | 36 | 1.0% | 27 | 1.7% |  |
|  | Type 2 | 323 | 19.7% | 729 | 20.1% | 329 | 20.1% |  |
| CKD** | None | 1296 | 79.0% | 2610 | 71.9% | 1205 | 73.8% | <0.001 |
|  | Mild | 296 | 18.0% | 898 | 24.7% | 347 | 21.2% |  |
|  | Moderate or Severe | 49 | 3.0% | 124 | 3.4% | 81 | 5.0% |  |

aLRTD, acute lower respiratory tract disease; CCI, Charlson comorbidity index; CKD, chronic kidney disease; COPD, chronic obstructive pulmonary disease; CRDE, chronic respiratory disease exacerbation; HF, heart failure; IHD, ischaemic heart disease; NP-LRTI, non-pneumonic lower respiratory tract infection; PCR, polymerase chain reaction; SD, standard deviation
† In the UK, patients aged ≥65 years are eligible for Pneumococcal vaccination (PneumoVax®, PPV23) once, and annual influenza vaccine
\* Hypertension was only included if causing other cardiac complications
\*\* Chronic kidney disease (CKD) was classified as mild if stage 1-3; moderate/severe if stage 4-5, end-stage renal failure or there was dialysis dependence

**Extended data 5:**
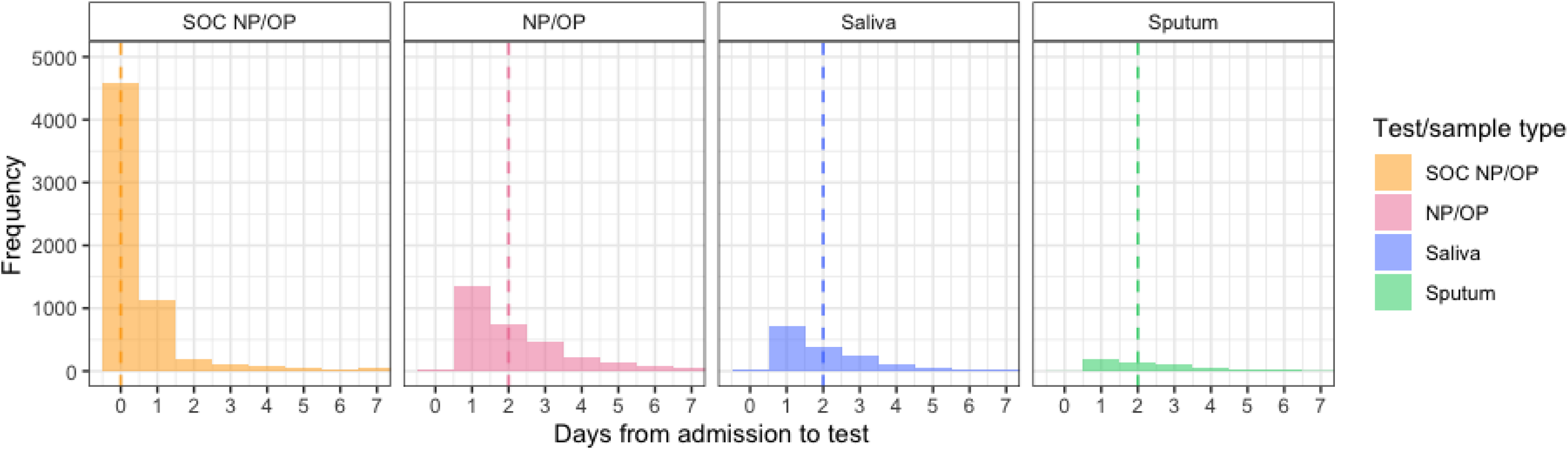
Duration of sampling following admission for aLRTD. The frequency of duration from admission to a participants first RSV test for a Standard of care (SOC NP/OP) test, or a research lab NP/OP sample, saliva sample or sputum sample. Only 0 – 7 days are shown. The dashed bars show the median number of days from admission the RSV test.

**Extended data 6:**
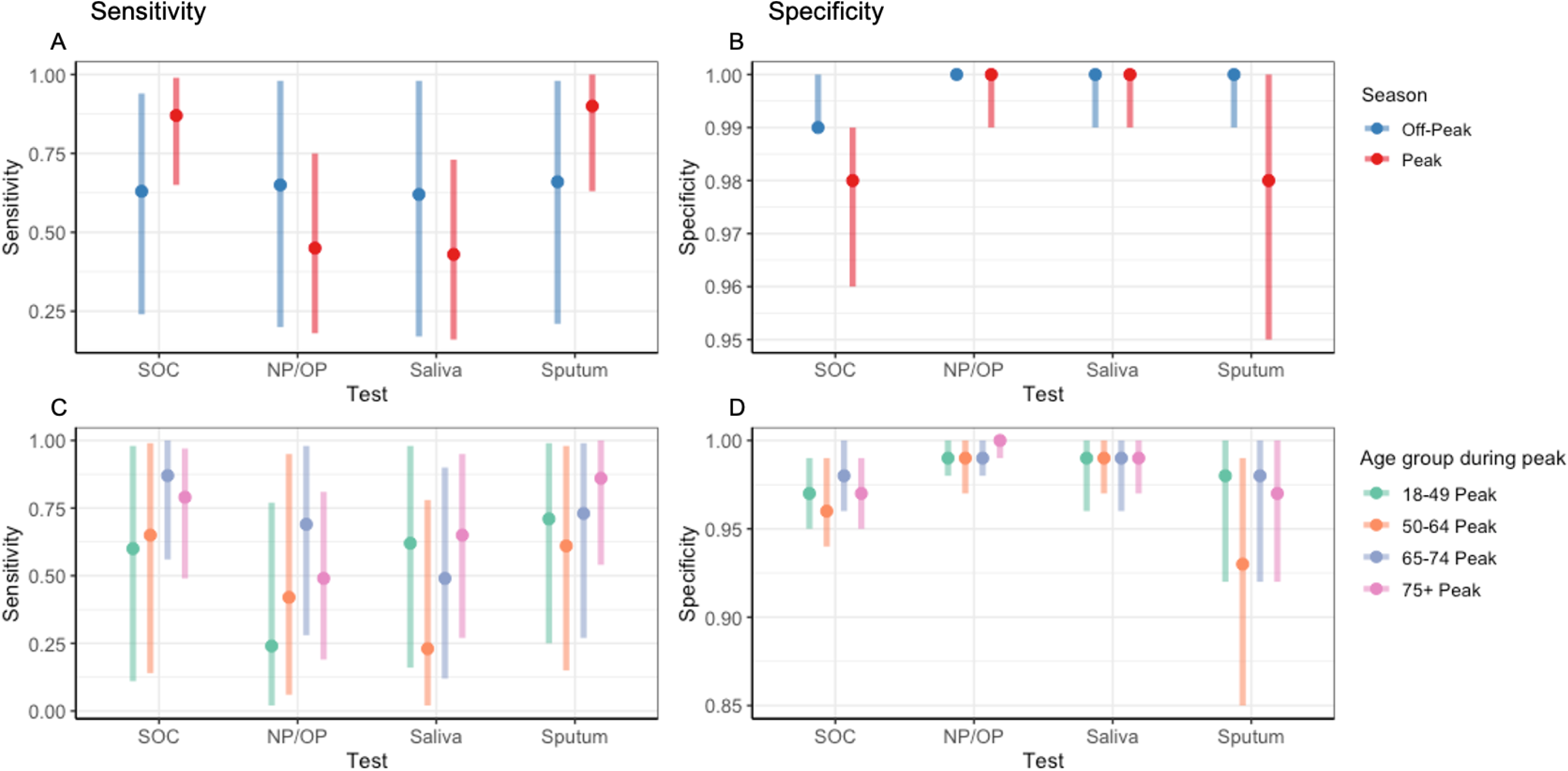
Test sensitivity and specificity inferences from a LC model run on different participant subgroups. Point estimates of inferences (mean) of test sensitivity (A/C) and specificity (B/D) from a latent class model accounting for test error and multiple testing run on different participant subgroups: Peak (Nov – Feb) and off-peak RSV season (A/B), and age groups during peak season (C/D). Bars show 95% credible intervals.

**Extended data 7:**
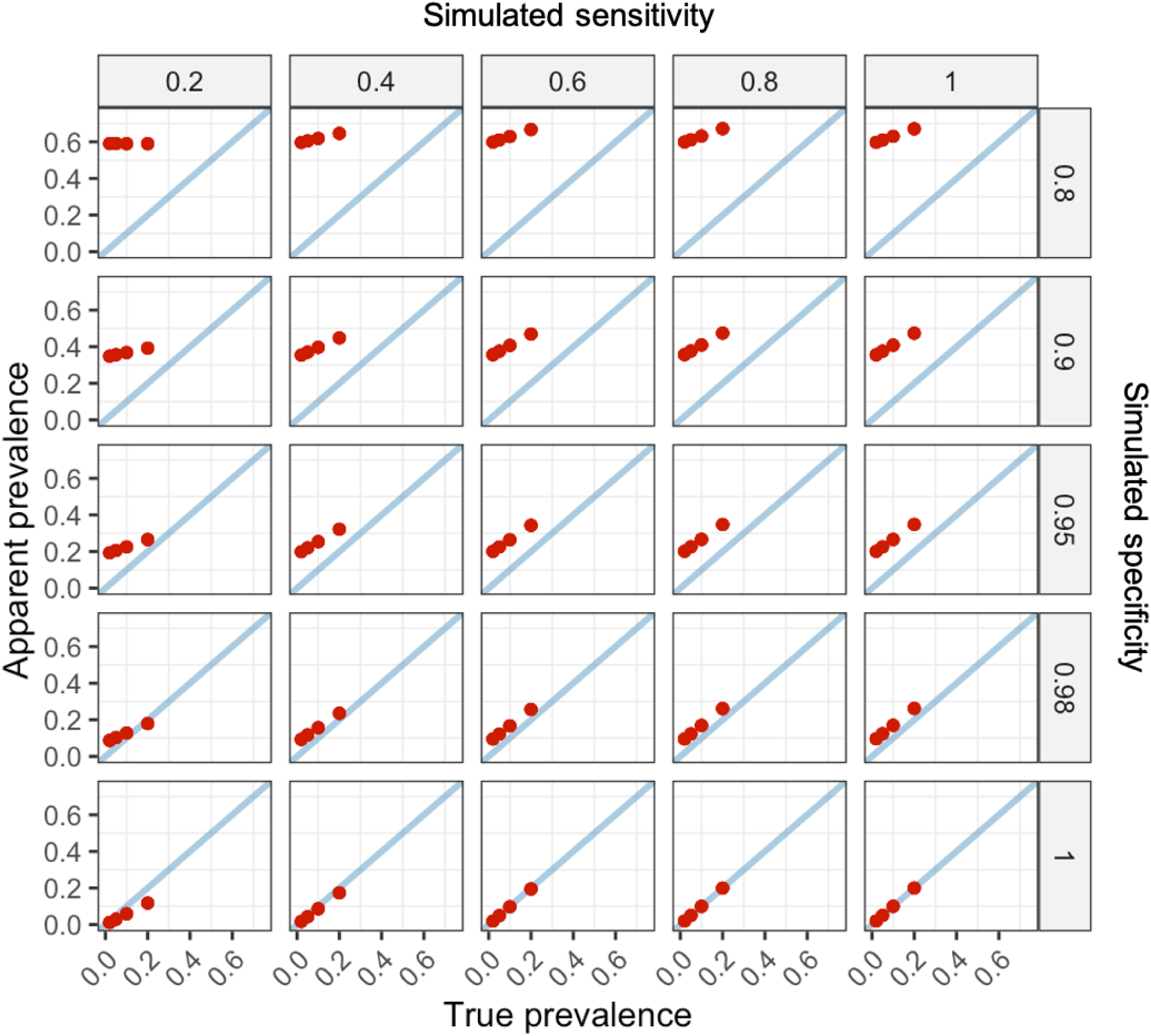
Relationship between apparent and true prevalence. The relationship between apparent/observed prevalence (test-positivity; y-axis) and true prevalence (x-axis) at different sensitivities and specificities from simulated data (N = 100000) with known parameter values. Apparent prevalence is based upon any of four tests with identical sensitivity and specificity being positive. Simulated sensitivity (top) and simulated specificity (right) values are the mean simulated values across the 4 tests.

**Extended data 8:**
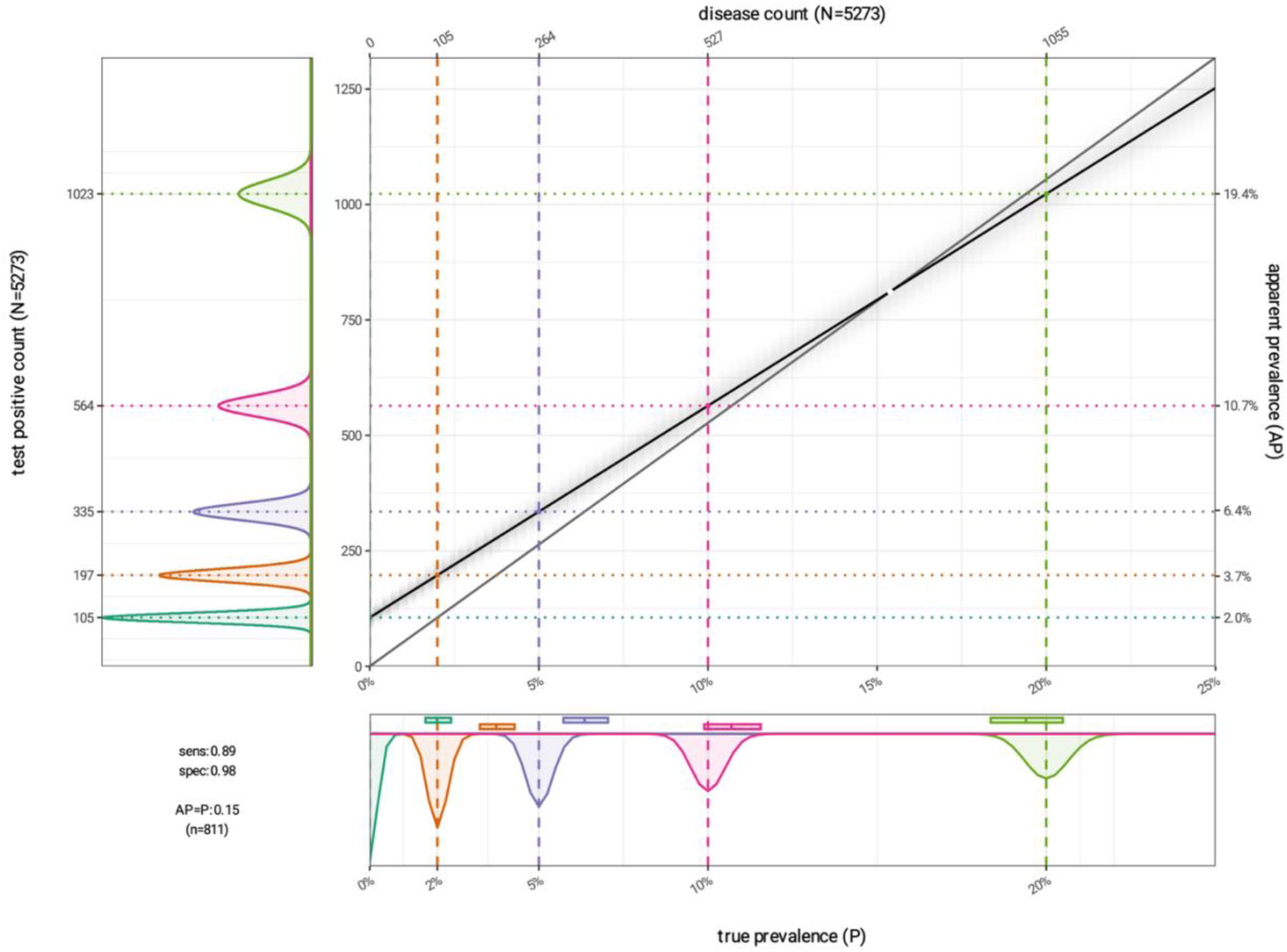
Rogan-Gladen estimator. A visualisation of the Rogan-Gladen estimator (Rogan and Gladen, 1978) showing the relationship between true prevalence (x-axis), number of test-positives (y-axis), apparent/observed prevalence (test-positivity; secondary y-axis), and number of disease positive individuals (secondary x-axis). The grey line represents perfect agreement between true and apparent prevalence, and the black line represents the relationship between apparent and true prevalence with test error. Sensitivity and specificity values are those inferred by a latent class model for the SOC test (Table 4), and the number of individuals is based on the number tested by SOC in the study (N= 5273). The dashed and dotted lines show the relationship between true and apparent prevalence, and test-positive and disease counts at different true prevalence values (0, 2, 5, 10, 20). The x and y plots show uncertainty around these estimates. The boxplots on the x plot show the apparent prevalence distribution for each true prevalence value. The white point represents the point at which apparent prevalence becomes an underestimation of true prevalence.

