## Supplementary Material for "Improving the accuracy of Respiratory Syncytial Virus (RSV) incidence among hospitalised adults in Bristol, UK"

### LATENT CLASS METHODS

The latent class model was built using a Bayesian framework in ‘Stan’, with NUTS (No-U-Turn Sampler) Hamiltonian Monte Carlo MCMC used to sample from the posterior distributions. The probability of an individual having a particular pattern of test outcomes is conditional on disease prevalence and test sensitivity and specificity, which are unknown latent parameters estimated by the model (from Keddie et al., 2023):

$$P(Y \mid \pi, Se, Sp) = \prod_{i=1}^{N}((\pi \prod_{j=1}^{R} Se_j^{y_{ij}}\,(1-Se_j)^{1-y_{ij}}\,) + ((1-\pi)\prod_{j=1}^{R} Sp_j^{(1-y_{ij})}(1-Sp_j)^{y_{ij}}\,))$$

$\pi = prevalence$

$y = binary\ test\ outcome$

$Y = pattern\ of\ multiple\ test\ outcomes$

$i = individual$

$N = total\ number\ of\ individuals$

$j = test$

$R = total\ number\ of\ tests$

Test dependence can be built into the model through inclusion of a random effect in the estimation of sensitivity and specificity of the research lab tests:

$Se_{ij} = g^{-1}\left(\alpha_{j1} + \beta_{j1}s_{i1}\right)$

$Sp_{ij} = g^{-1}\left(\alpha_{j0} + \beta_{j0}s_{i0}\right)$

$g^{-1} = link\ function$

$\alpha = test\ sensitivity\ or\ specificity$

$\beta = test\ dependence\ scaling\ factor$

$s = random\ effect$

The prior distribution for prevalence was defined as uniform between 0 and 1. The prior distributions for test sensitivity were defined as uniform between (1-specifcity) and 1 to ensure the probability of a positive test for an individual with disease is higher than for an individual without disease. The prior distributions for test specificity were defined as following a beta(10, 1) distribution, which assumes that there is a 95% probability of specificity being > 74%. This prior reflects the belief that specificity is skewed towards 1 without being overly constraining, and model inferences were not sensitive to changes in this prior (Supplementary, Data 5). When conditional dependence is included in the model between the research lab tests, the prior distribution for the individual-level random effect is defined as a normal distribution, and the random effect scaling parameters (beta0 and beta1) are defined as a gamma(1,1) distributions. This allows changes in the random effect to change the sensitivities of all research lab tests by the same amount, and the specificities of all research lab tests by the same amount, for each individual. The standard of care (SOC) test is considered independent.

**Multiple testing**

Test error associated with multiple testing was quantified by combining individual inferred test sensitivities and specificities in the 'generated quantities' block in Stan. As the apparent (observed) prevalence was based upon any positive test being considered RSV-positive and only all negative tests being considered RSV negative, combined test sensitivities and specificities for each test combination were calculated using the OR rule as follows (Zhou et al., 2002; Weinstein et al., 2005):

$$Overall\ test\ combination\ sensitivity = P(Y = 1 \mid d = 1) = 1 - \prod_{j=1}^{R} \left(1 - Se_j\right)$$

$$Overall\ test\ combination\ specificity = P(Y = 0 \mid d = 0) = \prod_{j=1}^{R} Sp_j$$

The positive predictive value (PPV), the proportion of positive test results that are true positives, and the negative predictive value (NPV), the proportion of negative test results that are true negatives, were calculated for each test combination as follows:

$$Overall\ test\ combination\ PPV = P(d = 1 \mid Y = 1) = \frac{\pi\left(1 - \prod_{j=1}^{R}\left(1 - Se_j\right)\right)}{\pi\left(1 - \prod_{j=1}^{R}\left(1 - Se_j\right)\right) + 1 - \pi\left(1 - \prod_{j=1}^{R} Sp_j\right)}$$

$$Overall\ test\ combination\ NPV = P(d = 0\ |Y = 0) = \frac{1 - \pi\left(\prod_{j=1}^{R} Sp_j\right)}{1 - \pi\left(\prod_{j=1}^{R} Sp_j\right) +\ \pi(\prod_{j=1}^{R}\left(1 - Se_j\right))}$$

Rogan-gladen estimation, which is an established method for estimating true prevalence from test results whilst accounting for imperfect sensitivity and specificity, was used to quantify the threshold between apparent prevalence being an over-estimation or an under-estimation of true prevalence at inferred test error rates (Rogan and Gladen, 1978):

$$True\ prevalence = \frac{\left(Apparent\ prevalence + (Sp - 1)\right)}{Sp + (Se - 1)}$$

### LATENT CLASS MODEL VALIDATION

Model performance was assessed using simulated data with known parameters of prevalence, test sensitivity and test specificity. Simulated data were generated as a population of individuals, *N*, which were randomly assigned a carriage status, positive or negative, with a known probability *p.* The true prevalence, therefore, has an expectation value of *Np/N*. The outcomes of a set of four tests *y* were then simulated on each individual in the population, generating a set of four test results based on the set sensitivity, specificity and testing-rate parameter for each test. These simulated test results were used to test the performance of the latent class models in their ability to infer test sensitivity, specificity and true prevalence accurately from datasets with known parameters.

**Supplementary, Data 1: Latent class inferences with test independence.** LC model, with assumed test independence, sensitivity and specificity inferences (mean with 95% credible intervals) from study data (N= 6906). The LC model inferred a prevalence of 1.9% (95%CI 1.3 – 2.7%).

The inclusion of test dependence between the research lab tests increased inferred SOC specificity from 0.97 to 0.98 and resulted in a slightly higher inferred prevalence (2.3% (95%CI 1.4-3.7%); (Table 3). This is to be expected when correlations between the research lab tests are not accounted for, as given a true positive SOC test, false negative results in all three research tests will act to push estimates of the specificity of the SOC test downwards more if the tests are considered completely independent than if they are considered to be three correlated observations of a patient whose RSV has resolved.

| Test | Sens | Spec |
|---|---|---|
| SOC | 0.89 (0.76-0.98) | 0.97 (0.97-0.98) |
| NP | 0.57 (0.37-0.77) | 1.00 (1.00-1.00) |
| Saliva | 0.55 (0.33-0.79) | 1.00 (1.00-1.00) |
| Sputum | 0.90 (0.69-1.00) | 0.99 (0.98-1.00) |

**Supplementary, Data 2: Latent class with test independence Vs simulated data.**

LC model inferences of a) prevalence, b) sensitivity and c) specificity (mean across tests) compared to simulated values when run with simulated RSV test data (N = 1000) at different simulated prevalences, sensitivities and specificities. Tests are assumed to be independent, the simulated sensitivity and specificity are the same for all 4 tests, and the testing-rate is assumed to be 100% for every test. 95% credible intervals are shown. The orange boxes show results from simulated data within parameter ranges expected in this study. If no points are shown, then simulations were not performed for this parameter combination.

a) Prevalence

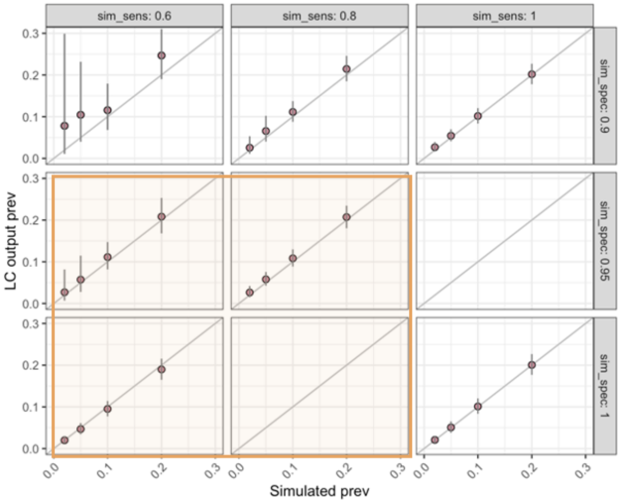


b) Sensitivity

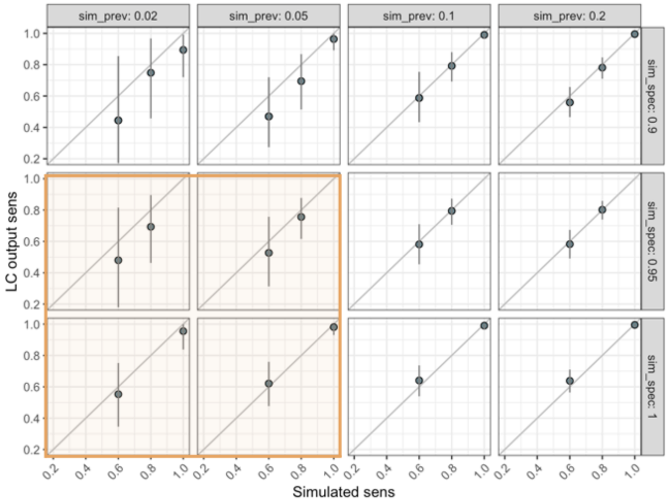


a) Specificity

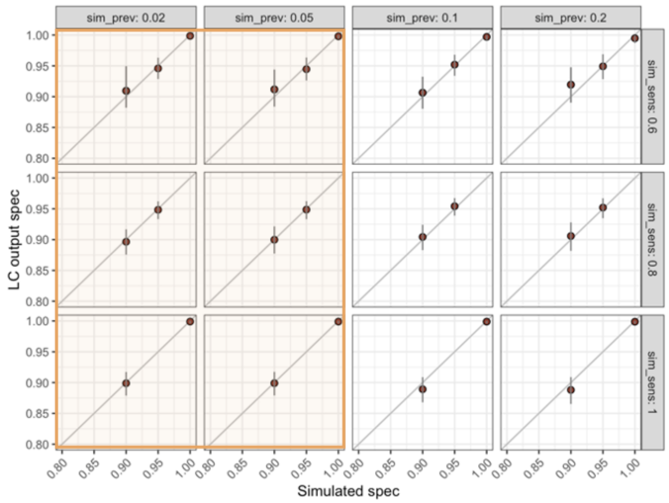

**Supplementary, Data 3: Latent class with test independence Vs simulated data with variable test parameters.**

LC model with assumed test independence, sensitivity (a) and specificity (b) mean inferences compared to simulated parameters. Data were simulated at variable test sensitivity, specificity and testing-rates based on study data testing-rates and model inferences using the study data (Supplementary, Data 1). 95% credible intervals are shown. The LC model recovered the simulated prevalence of 1.9% (1.4 - 2.9).

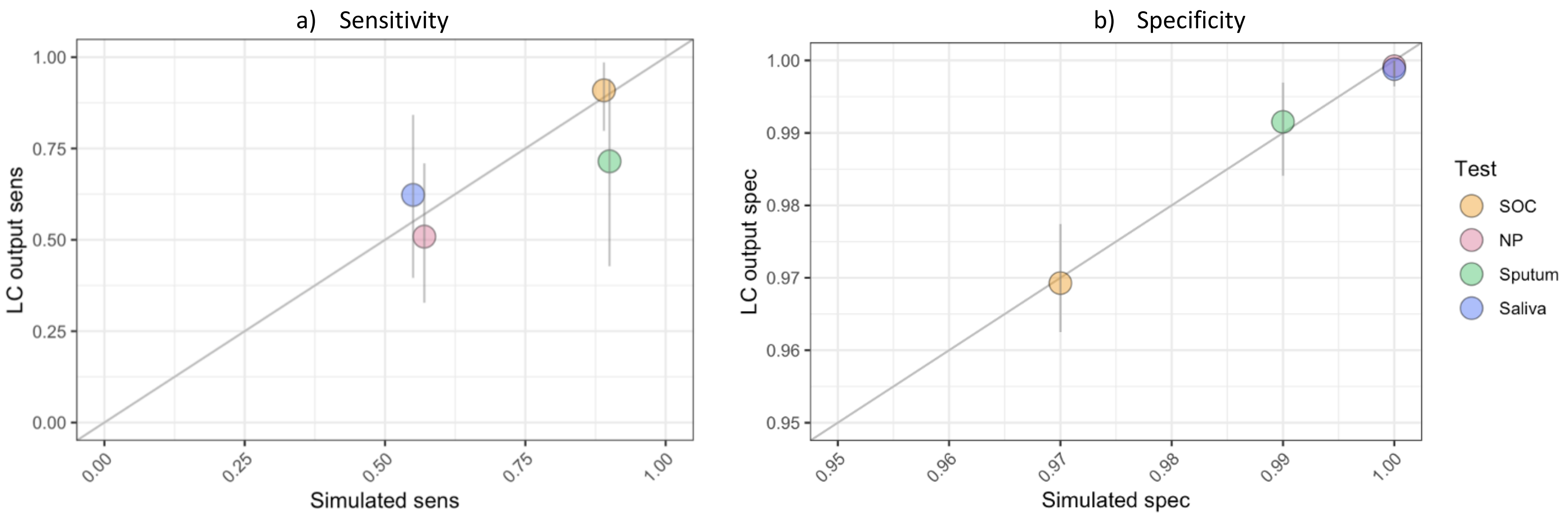

**Supplementary, Data 4: Latent class with test dependence Vs simulated data with variable test parameters.** LC model with research lab test dependence, prevalence (a), sensitivity (b) and specificity (c) inferences compared to simulated parameters. Data were simulated (N=5000) at different test sensitivities and specificities for the SOC vs research lab tests, at a true prevalence of 0.02 and 0.05. The simulated sensitives and specificities were the same for the three research lab tests to introduce test correlation into the simulated dataset. Individual test testing-rates were based on study data. 95% credible intervals are shown.

a) Prevalence

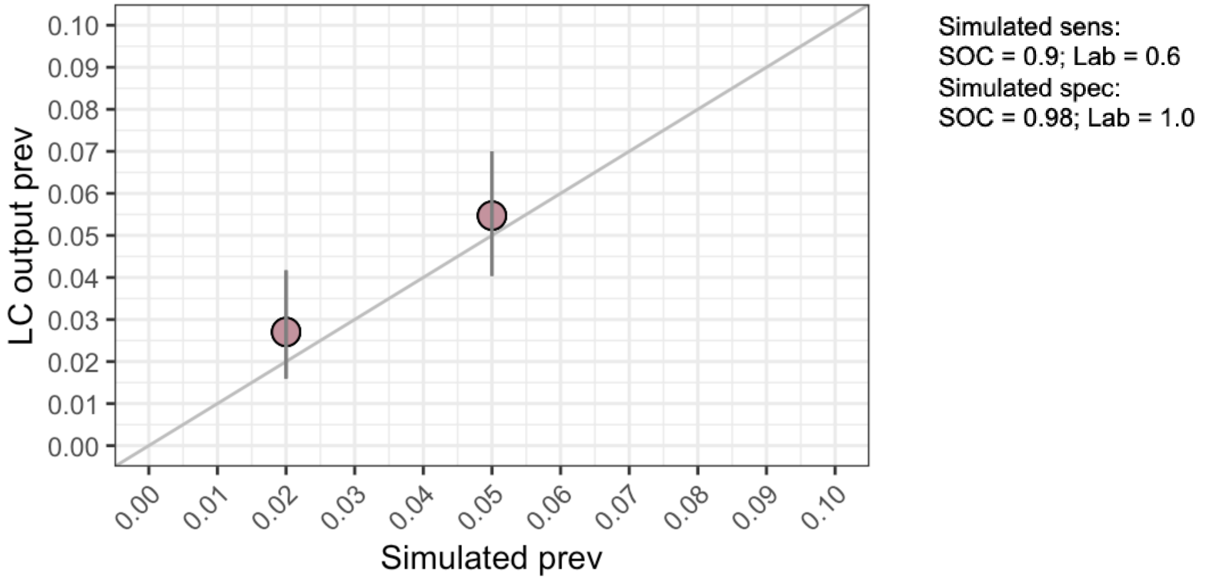


b) Sensitivity

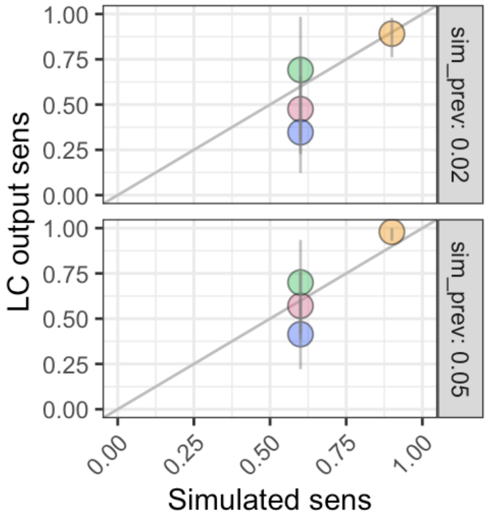


c) Specificity

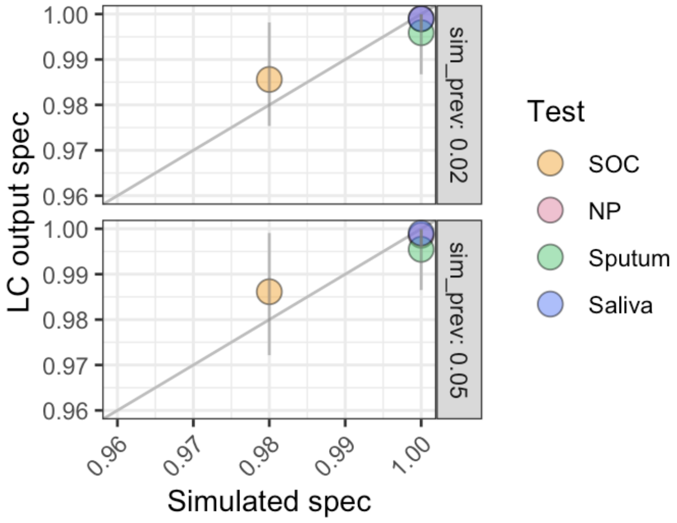

**Supplementary, Data 5. Sensitivity analysis of priors for test specificity.** Shows prior descriptions and inferred LC model parameters. 95%CI values are shown in brackets. NA values indicate that the priors did not fit the data.

| **Priors for specificity** | **Prior description** | **Inferred specificity** | | | | **Inferred prevalence (%)** |
|---|---|---|---|---|---|---|
| | | **SOC NP/OP** | **Lab NP/OP** | **Lab Saliva** | **Lab Sputum** | |
| **Beta(1, 1)** | Flat, non-specific prior.<br>>95% chance value is >0.05; mean = 0.5 | 0.984<br>(0.979 - 0.991) | 0.999<br>(0.998 – 1.00) | 0.999<br>(0.997 – 1.00) | 0.993<br>(0.982 – 0.999) | 2.3 (1.4 - 3.6) |
| **Beta(10, 1)** | >95% chance value is >0.741; mean = 0.909 | 0.984<br>(0.979 – 0.991) | 0.999<br>(0.998 – 1.00) | 0.999<br>(0.997 – 1.00) | 0.993<br>(0.983 – 0.999) | 2.3 (1.4 - 3.7) |
| **Beta(82, 1)** | >95% chance value is >0.964; mean = 0.988 | 0.985<br>(0.979 – 0.993) | 0.999<br>(0.998 – 1.00) | 0.999<br>(0.997 – 1.00) | 0.994<br>(0.985 – 0.999) | 2.4 (1.4 - 3.9) |
| **Beta(215, 1)** | Highly specific prior skewed to 1.0.<br>>95% chance value is >0.986; mean = 0.995 | NA | NA | NA | NA | NA |

**Supplementary, Data 6: Latent class model inferences with different SOC test types.**

LC model test sensitivity and specificity inferences (mean with 95% credible intervals) from study data (N= 6906), with standard of care tests included as the three different SOC panel tests used (abbreviated here to 'SOCpanel', SOCbiofire', 'SOCquad'). Only a participants first RSV test was included, and the apparent (observed) prevalence was 3.5% (95%CI 3.1 – 4.0%; 242/6906). Dependence was modelled between the 3 lab tests and between the 3 SOC tests. The LC model inferred a prevalence of 1.9% (95%CI 1.3 – 2.7%).

| Test | Sens | Spec |
|---|---|---|
| SOCpanel | 0.93 (0.73-1.00) | 0.97 (0.96-0.99) |
| SOCbiofire | 0.78 (0.48-0.97) | 0.99 (0.98-1.00) |
| SOCquad | 0.57 (0.20-0.92) | 0.98 (0.96-0.99) |
| NP | 0.47 (0.17-0.75) | 1.00 (1.00-1.00) |
| Saliva | 0.44 (0.14-0.75) | 1.00 (1.00-1.00) |
| Sputum | 0.85 (0.53-0.99) | 0.99 (0.98-1.00) |

**Supplementary, Data 7: Subgroup test-positivity vs latent class inferred prevalence, sensitivity and specificity from a LC model with different SOC test types.**

Inferences of prevalence (A), sensitivity (B) and specificity (C) from a latent class model accounting for test error and multiple testing run on different participant subgroups. A) Point estimates of test-positivity (any positive / tested; grey) and inferred true prevalence (mean; pink), run for the subgroups peak season (Nov-Feb), off-peak season, and age groups during the peak season. Bars show 95% CI's (grey: Clopper and Pearson confidence intervals; pink: credible interval). B/C) Point estimates of sensitivity and specificity (mean) for different age groups during the peak season (Nov-Feb). Bars show 95% credible intervals.

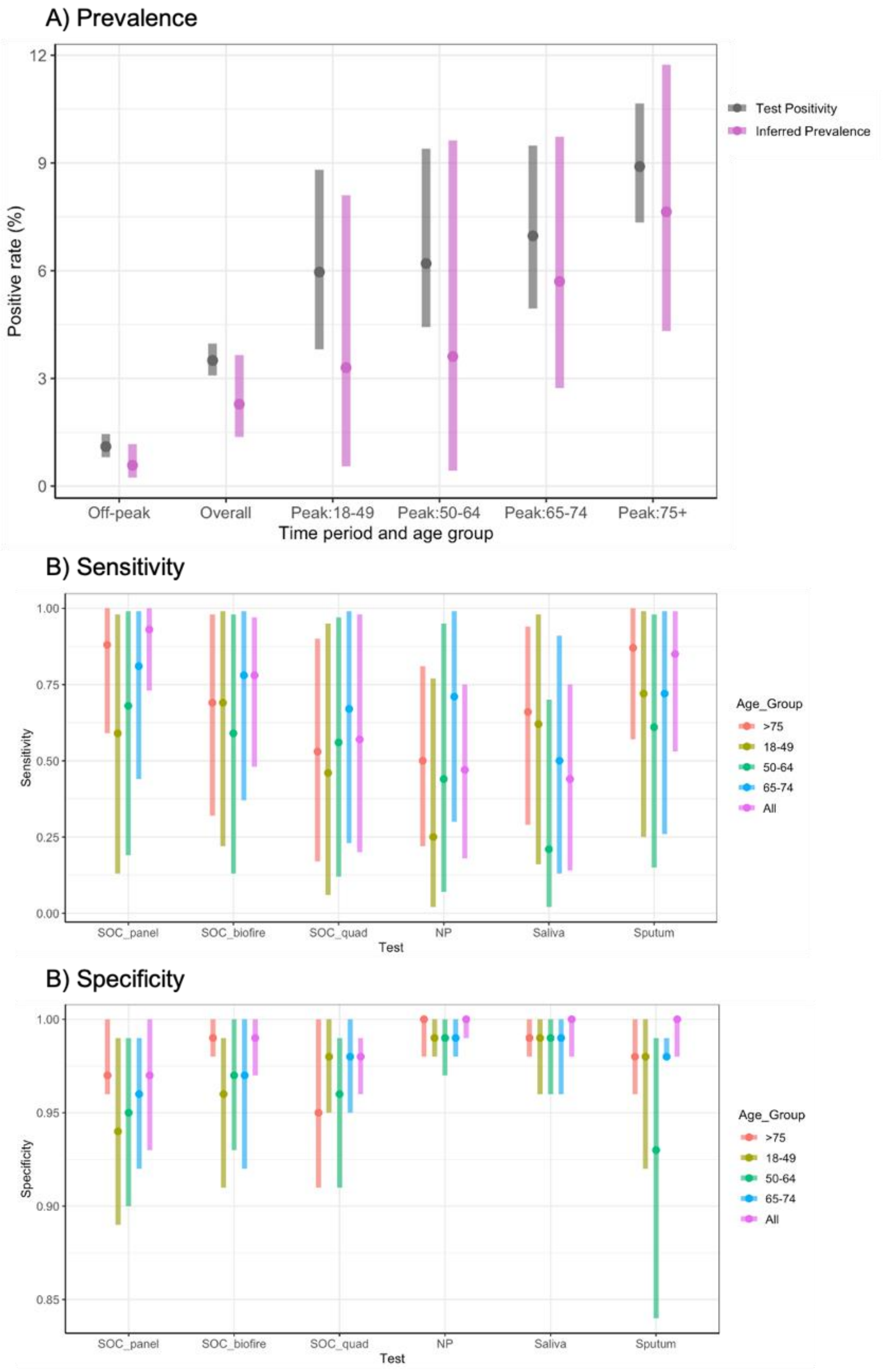
